# Preoperative Social Connection and Postoperative Outcomes in Adults Undergoing Surgery: A Systematic Review and Meta-analysis

**DOI:** 10.64898/2026.09.15.26363083

**Authors:** Chuan Yin, Youhao Wang, Long Jiang, Xihao Huang, Zhicheng Zhang, Zehao Jing

**Author notes:** **Corresponding authors:** Chuan Yin, MD, PhD; Department of Orthopaedic Surgery, Shanghai Ninth People’s Hospital, Shanghai Jiao Tong University School of Medicine, 639 Zhizaoju Road, Huangpu District, Shanghai 200011, China; **Email:**. Zehao Jing, MD, PhD; Department of Orthopaedics, Peking University Third Hospital, 49 North Garden Road, Haidian District, Beijing 100191, China; **Email:**. Chuan Yin and Youhao Wang contributed equally as co–first authors.

## Abstract

**Importance:** Social connection is routinely recorded in surgical cohorts but rarely analyzed as an exposure, leaving its prognostic value unsummarized.

**Objective:** To determine whether weaker preoperative social connection is associated with adverse postoperative outcomes.

**Data Sources:** PubMed/MEDLINE, Embase, Scopus, Web of Science, CINAHL, PsycInfo, CENTRAL; inception to July 17, 2026; 33,301 records, 20,399 after deduplication.

**Study Selection:** Studies of adults undergoing surgery with preoperatively ascertained social connection analyzed against a protocol-listed outcome; of 1,550 reports assessed, 454 reports of 445 studies were included.

**Data Extraction and Synthesis:** PRISMA and MOOSE reporting; PROSPERO CRD420261449181. Estimate selection followed a frozen, direction-blind hierarchy; k denotes analysis weight units. Random-effects models used restricted maximum likelihood (REML) with modified Hartung-Knapp intervals; risk of bias, result-level Quality in Prognosis Studies (QUIPS); certainty, prognostic Grading of Recommendations Assessment, Development and Evaluation (GRADE). Artificial intelligence agents screened and extracted; 2 reviewers unblinded to their output confirmed all 39 principal-analysis estimate records.

**Main Outcomes and Measures:** Postoperative all-cause survival (primary); early postoperative mortality, non-home discharge, unplanned readmission, any or major complications (secondary).

**Results:** Poolable estimates came from 72 of 445 studies, mostly of marital status. Each principal analysis included up to 63,779 to 298,340 patients or procedures (upper bounds; cohorts recur across outcomes, so no overall total). For the primary outcome, postoperative survival, the confidence interval (CI) included 1 (9 units; hazard ratio, 1.37; 95% CI, 0.99-1.89; certainty very low). Weaker connection was associated with early postoperative mortality (8 units; odds ratio, 1.50; 95% CI, 1.12-2.01; certainty low) and non-home discharge (9 units; OR, 1.95; 95% CI, 1.35-2.81; certainty low). CIs included 1 for unplanned readmission (9 units; OR, 1.14; 95% CI, 0.98-1.33; certainty low) and complications (4 units; OR, 1.10; 95% CI, 0.96-1.25; certainty very low). All estimable prediction intervals included the null. No eligible study estimated the association with failure to rescue.

**Conclusions and Relevance:** Preoperative social connection is a potential prognostic signal with substantial uncertainty. The largest association concerned discharge destination, partly shaped by the exposure itself, supporting early identification of support needs, not candidacy restriction. Whether changing social connection alters outcomes was not directly evaluated.

**Key Points:** *Question:* Among adults undergoing surgery, is weaker preoperative social connection associated with adverse postoperative outcomes?

*Findings:* In this systematic review and meta-analysis of 445 studies (33 in 5 principal analyses), all pooled estimates exceeded 1, the primary CI included 1 (HR, 1.37; 95% CI, 0.99-1.89) and CIs excluded 1 for early postoperative mortality (OR, 1.50) and non-home discharge (OR, 1.95), yet prediction intervals included 1 and certainty was low or very low.

*Meaning:* Preoperative social connection—in this literature, mostly marital status—is a potential prognostic signal with substantial uncertainty; it can inform support assessment and discharge planning, not candidacy.

## Introduction

Social connection—the ties linking a person to a partner, household, network and practical help—is routinely recorded in surgical cohorts, usually as a covariate, not the exposure.^1^ In 2023 the US Surgeon General made social connection a public-health priority.^2^

Surgery is a plausible setting for it to carry prognostic information: recovery happens where the surgical team is absent, and someone must manage wounds and medications, notice deterioration and seek help.^3,4^ Guideline-based preoperative assessment already asks surgeons to determine the patient’s social support system.^5^

The surgical literature nevertheless supports no dependable summary: reports differ in construct, outcome definition, time origin and adjustment, and many of the largest series are registry analyses sharing patients, so publication and participant counts overstate the independent evidence, and pooling across incompatible definitions manufactures unsupported precision.

We conducted a systematic review and meta-analysis (PROSPERO CRD420261449181), pooling one umbrella contrast—weaker versus stronger connection—with each study’s construct carried as a frozen axis, and report average associations, their certainty, and what this literature cannot yet answer.

## Methods

### Registration and reporting

The review follows Preferred Reporting Items for Systematic Reviews and Meta-Analyses (PRISMA) 2020 and Meta-analysis of Observational Studies in Epidemiology (MOOSE) guidance.^6,7^ The analysis plan, exposure ontology, outcome definitions and analysis-level rules were frozen before synthesis; departures from the registered protocol are itemized in eTable 1. This synthesis of published aggregate data involved no human participants and required no ethics review.

### Data Sources and Searches

We searched 7 databases (PubMed/MEDLINE, Embase, Scopus, Web of Science, CINAHL, PsycInfo, CENTRAL) from inception to July 17, 2026, without language limits, yielding 33,301 records and 20,399 after deduplication (eMethods 1). Registered citation searching was not performed (eTable 1).

### Selection Criteria

Outcomes followed a frozen controlled vocabulary, with time origin and window carried per estimate rather than fixed by the outcome label (eMethods 2). We included studies of adults undergoing surgery in which social connection was ascertained preoperatively and analyzed against a protocol-listed outcome, and excluded those in which the exposure appeared only as a baseline characteristic or unreported covariate, social connection was an intervention, or only outcomes beyond 1 year were reported.

### Exposure Definition

Eligible exposures were mapped onto a frozen hierarchy (structural: marital or partnership status, living arrangement, network size and isolation; functional: perceived or functional support, caregiver availability; no eligible measure captured relationship quality), with unmatched measures left unclassified. The principal families pool the *umbrella* contrast—weaker versus stronger connection, whichever construct a study measured (Table 1; a documented departure from registered pooling, eTable 1). No nonmarital construct contributed more than 2 units to any principal family (eTable 2), and a post hoc marital-restricted sensitivity analysis is reported (eTable 3).

**Table 1.** Evidence Base and Characteristics of the Five Principal Analyses and the Broad All-Origin Survival Sensitivity Node.

| Analysis | Analysis type | k (analysis weight units) | No. of studies | Dependence clusters | Publication years | Surgical specialties (studies) | Analyzed contrast (units) | Analyzed patients, procedures or discharges (upper bound) |
| --- | --- | --- | --- | --- | --- | --- | --- | --- |
| Postoperative all-cause survival (primary) | Primary | 9 | 9 | 9 | 1998-2022 | 4 Transplantation; 3 Cardiac; 2 Orthopaedic | 5 Marital/partnership; 2 Perceived/functional support; 1 Living arrangement; 1 Other/unclassified | ≤127,991 (136-96,764 per unit) |
| Broad all-origin survival (maximum-coverage sensitivity) | Sensitivity | 16 | 16 | 16 | 1998-2025 | 5 Transplantation; 4 Gastrointestinal/oncologic; 4 Cardiac; 3 Orthopaedic | 11 Marital/partnership; 2 Perceived/functional support; 2 Living arrangement; 1 Other/unclassified | ≤299,735 in the 15 of 16 units with a reported size |
| Early postoperative mortality | Key secondary | 8 | 8 | 8 | 2010-2026 | 4 Mixed/other surgical; 2 Gastrointestinal/oncologic; 1 Cardiac; 1 Orthopaedic | 6 Marital/partnership; 1 Living arrangement; 1 Network/isolation | ≤63,779 (382-27,905 per unit) |
| Non-home discharge | Key secondary | 9 | 9 | 9 | 2011-2025 | 5 Orthopaedic; 2 Gastrointestinal/oncologic; 1 Vascular; 1 Cardiac | 8 Marital/partnership; 1 Living arrangement | ≤129,988 (135-106,752 per unit) |
| Unplanned readmission across all eligible windows | Key secondary | 9 | 9 | 9 | 2008-2025 | 6 Orthopaedic; 2 Gastrointestinal/oncologic; 1 Cardiac | 7 Marital/partnership; 2 Living arrangement | ≤298,340 (175-167,265 per unit) |
| Any/major postoperative complications | Key secondary | 4 | 4 | 4 | 2019-2025 | 4 Gastrointestinal/oncologic | 3 Marital/partnership; 1 Living arrangement | ≤148,525 (151-106,752 per unit) |

### Units of Analysis

We report 5 nested quantities: *reports* (publications), *studies*, *cohort entities* (source-defined cohorts after collapsing duplicate reporting), *dependence clusters* (cohort entities sharing participants under registry-provenance rules), *analysis weight units* (one per cluster, receiving weight in a model; eMethods 2). Each outcome-specific pooled analysis is a *family* (Table 1); throughout, **k denotes analysis weight units**, not reports or studies, and is not additive across families.

### Study Selection and Data Extraction

Review tasks were executed by artificial intelligence (AI) agents (Claude Code, Anthropic; eMethods 3, with access terms for texts entered) under numbered written investigator directives and hash-frozen specifications; investigators ruled on protocol ambiguities and authorized the analytic release. Titles and abstracts were screened by 2 agents blinded to each other (κ = 0.87, agent-to-agent concordance, not accuracy against a human standard); 598 disagreements were adjudicated by a third agent. Full texts were assessed likewise, a verifier agent re-examining a risk-weighted sample. Agents extracted with a source locator for every analyzed estimate. The 39 estimate records (from 33 studies) underlying the principal analyses were confirmed by 2 people answering 312 field-level questions against the printed sources, separately but seeing the same AI-proposed values (not blinded independent dual review; eMethods 4); 0 corrections resulted. Analyzed sample size was not among those fields; sizes not established at extraction were revisited after the lock by one agent with independent re-derivation by a second, then checked against the printed source by 1 of 2 investigators (18 sizes, 10 not-printed rulings; 0 corrections); 9 later-recovered bounds and 18 locator-checked cohort totals were not. Where the fitted model’s size was never printed, the tightest printed count bounding it from above (arm sum, analyzed cohort or enclosing cohort) is reported and marked ≤ (exact when an arm sum exhausts the analyzed set); one estimate remains unsized (eMethods 5).

### Estimate Selection

Where a study reported several eligible estimates for one outcome, one entered each analysis through a frozen, source-based hierarchy—exposure contrast, outcome definition and time origin first, then adjustment stratum, model type and reporting completeness—that never used effect direction, interval width or null-crossing. Every legal alternative was re-run as a multiverse analysis (eTable 4).

Contributing estimates mixed unadjusted, baseline-adjusted, post-exposure-conditioned and mixed or unclear models, a frozen per-estimate axis (Table 2).

**Table 2.** Summary of Findings.

| Outcome | Analysis type | k (analysis weight units) | No. of studies | Effect measure | Pooled estimate | 95% CI | I<sup>2</sup> | τ<sup>2</sup> | 95% prediction interval | Adjustment basis of contributing estimates | Results at high risk of bias, % of random-effects weight | GRADE certainty (Claim B) |
| --- | --- | --- | --- | --- | --- | --- | --- | --- | --- | --- | --- | --- |
| Postoperative all-cause survival (primary) | Primary | 9 | 9 | HR | 1.37 | 0.99-1.89 | 74.1 % | 0.107 | 0.59-3.17 | 1 mixed/unclear; 1 post-exposure-conditioned; 4 baseline-adjusted; 3 unadjusted | 77.72 | Very low |
| Broad all-origin survival (maximum-coverage sensitivity) | Sensitivity | 16 | 16 | HR | 1.23 | 0.995-1.516 | 76.4 % | 0.071 | 0.67-2.26 | 2 mixed/unclear; 3 post-exposure-conditioned; 5 baseline-adjusted; 6 unadjusted | 70.31 | Very low |
| Early postoperative mortality | Key secondary | 8 | 8 | OR | 1.50 | 1.12-2.01 | 66.8 % | 0.039 | 0.85-2.65 | 4 post-exposure-conditioned; 3 baseline-adjusted; 1 unadjusted | 51.93 | Low |
| Non-home discharge | Key secondary | 9 | 9 | OR | 1.95 | 1.35-2.81 | 78.1 % | 0.156 | 0.71-5.34 | 2 mixed/unclear; 4 post-exposure-conditioned; 2 baseline-adjusted; 1 unadjusted | 50.79 | Low |
| Unplanned readmission across all eligible windows | Key secondary | 9 | 9 | OR | 1.14 | 0.98-1.33 | 43.7 % | 0.013 | 0.84-1.56 | 1 mixed/unclear; 1 post-exposure-conditioned; 4 baseline-adjusted; 3 unadjusted | 63.16 | Low |
| Any/major postoperative complications | Key secondary | 4 | 4 | OR | 1.10 | 0.96-1.25 | 26.2 % | 0.003 | Not reported (k<5) | 1 mixed/unclear; 3 unadjusted | 100.0 | Very low |

### Statistical Analysis

Estimates were analyzed on the log scale, with standard errors from printed confidence limits, inverted where indexed on the stronger-connection group, as registered (eMethods 2). Random-effects models were fitted by restricted maximum likelihood on the model scale^8^, and confidence intervals (CIs) used a modified Hartung-Knapp adjustment (variance inflator floored at 1; eTable 4).^9,10^ Heterogeneity is summarized as IZ^11^ and, where k≥5, as a 95% prediction interval (not reported below).^12,13^ Odds ratios (ORs), hazard ratios (HRs) and risk ratios were never pooled together. Models were fitted by deterministic project code in Python 3.9.6 (NumPy 2.0.2, SciPy 1.13.1), reproduced by a separately written engine and independently in R 4.6.1 (eMethods 2). Leave-one-out is reported for every principal analysis and the multiverse wherever legal alternatives existed; funnel-plot asymmetry was examined exploratorily where k≥10. Two thresholds were frozen before synthesis: moderators or meta-regression only where k≥10, and ≥5 independent units sharing one window for a strict-window readmission analysis. Absolute risk differences and E-values^14^ were derived post hoc for those 2 associations.

### Risk of Bias and Certainty of Evidence

Risk of bias was assessed with the Quality in Prognosis Studies (QUIPS) tool at the level of the *result*, since one study can contribute results of differing quality.^15,16^ Certainty used the Grading of Recommendations Assessment, Development and Evaluation (GRADE) approach for prognostic factor evidence across five domains, starting at high without automatic downgrading for observational design.^17^ Study-level assessments were made in duplicate with third-assessor adjudication (assessor identifiers, eMethods 3); result-level judgments were derived by frozen rule and certainty computed from the frozen protocol rather than rated de novo, with indirectness not serious wherever exposure and outcome matched the registered definitions. Grades apply to the pooled effect claim only; evidence-map completeness is ungraded, and whether improving social connection would change outcomes is **not directly evaluated**.

### Outcomes

All principal outcomes are on the registered outcome list (primary designation reassigned before synthesis; eTable 1). Postoperative survival comprises a *strict primary node* (clock starting at the index operation or equivalent) and a *broad all-origin sensitivity node* admitting any time origin, reported for coverage only.

## Results

### Search and evidence landscape

Of 20,399 unique records screened, 18,420 were excluded at title/abstract and 1,979 reports sought; 252 were not retrieved, 177 failed full-assessment requirements, 1,550 were assessed. In all, 970 were excluded with reasons, 102 lacked evidence to classify and 24 carried terminal protocol-ambiguity states (eTable 5). Altogether, 454 reports of 445 studies were included, spanning 441 cohort entities and 421 dependence clusters (Figure 1; eMethods 2). Three corpus descriptions follow, not nested; eligible estimates spanned four overlapping domains: survival (100 studies), recovery failure (76), healthcare burden (172) and functional recovery (121; eFigure 1). At the study level, 127 studies met eligibility for the primary survival question (75 with a quantitative estimate, 52 direction-only). At the synthesis level, 72 studies (from all outcome families, not only the survival-eligible set) contributed to at least one fitted synthesis (62 dependence clusters; 216 analysis weight units, a union not a sum); 36 further clusters gave direction only.

**Figure 1.**
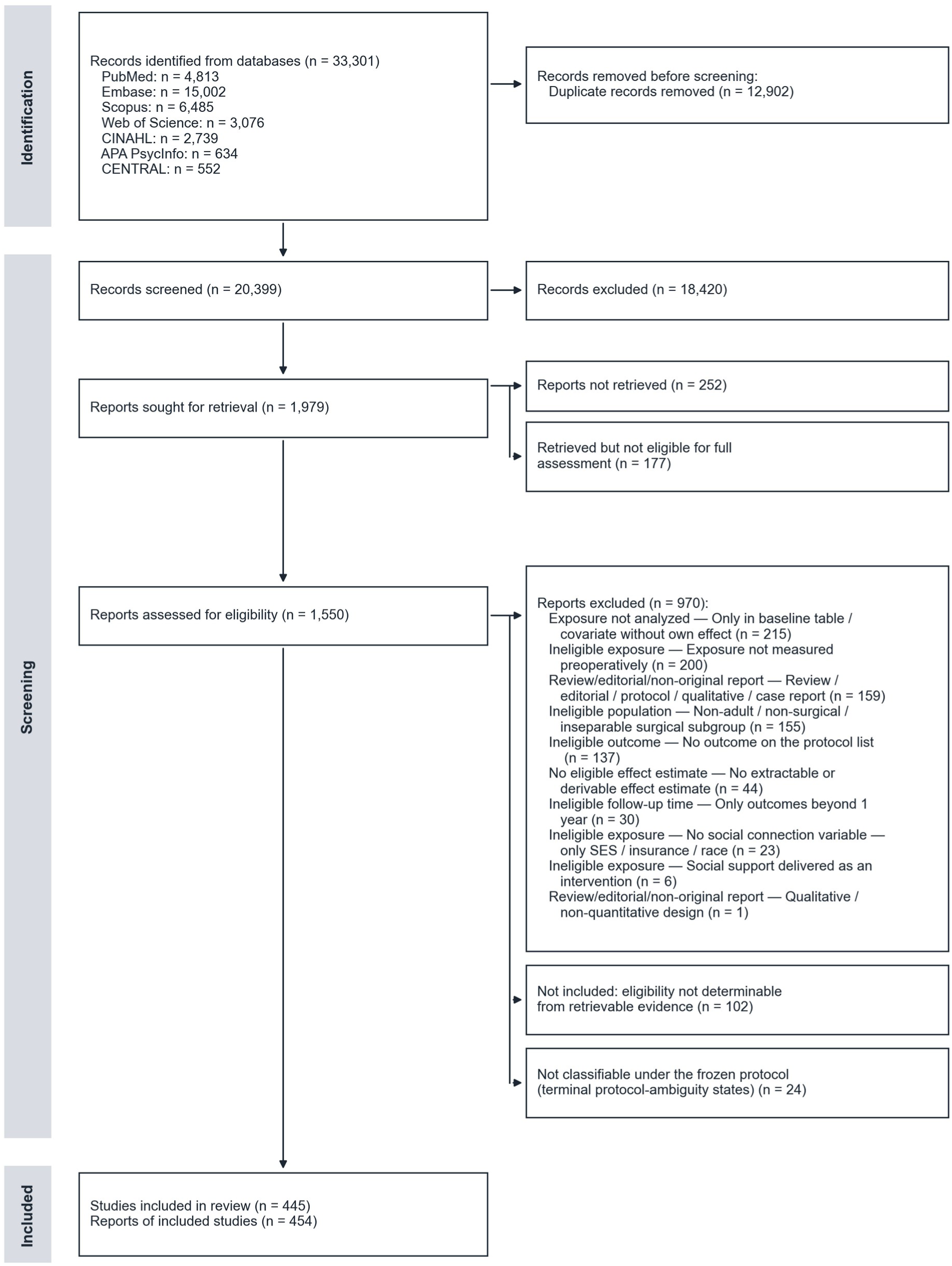
Flow of Records Through Identification, Screening and Inclusion (PRISMA 2020). Of 33,301 records identified from 7 databases, 12,902 duplicates were removed and 20,399 records were screened; 18,420 were excluded at title and abstract. Of 1,979 reports sought for retrieval, 252 were not retrieved and 177 were retrieved but not eligible for full assessment; 1,550 reports were assessed for eligibility, of which 970 were excluded with reasons (eTable 5), 102 could not have eligibility determined from retrievable evidence, and 24 could not be classified under the frozen protocol (terminal protocol-ambiguity states). 454 reports of 445 studies were included. All counts reconcile to eTable 5. ‘Retrieved but not eligible for full assessment’ denotes retrieved files that did not meet the locked requirements for entering full assessment (for example, abstract-only records or files that failed identity verification); ‘Not included: eligibility not determinable from retrievable evidence’ denotes reports whose retrievable evidence was insufficient to decide eligibility; ‘Not classifiable under the frozen protocol’ denotes terminal protocol-ambiguity states that were neither included nor excluded. CENTRAL, Cochrane Central Register of Controlled Trials; CINAHL, Cumulative Index to Nursing and Allied Health Literature; PRISMA, Preferred Reporting Items for Systematic Reviews and Meta-Analyses; SES, socioeconomic status.

### Exposure and study characteristics

Marital or partnership status was the analyzed contrast in 3 to 8 units of every principal analysis; living arrangement, perceived support, network measures and 1 unclassified loneliness item supplied the rest (Table 1; eTable 6; eTable 2). The 39 unique studies contributing to the principal and sensitivity analyses^18–56^ were concentrated in transplantation, cardiac, orthopedic and gastrointestinal or oncologic surgery (eFigure 1C; Table 1). Analyzed sizes were established for 45 of 46 contributing estimates (eTable 2); every principal analysis has a count, from ≤63,779 patients or procedures for early mortality to ≤298,340 for readmission (Table 1). Totals are upper bounds (sources usually print a cohort, not a model size), additive within a family only. Counts do not index precision: interval width is governed by between-unit variance and k; the largest unit supplied 76% of the primary survival count but 16% of its weight (82% and 14% for non-home discharge).

### Postoperative survival (primary; strict time origin)

Across 9 analysis weight units from 9 studies (eFigure 2; Figure 2A), the pooled estimate was above 1 (HR, 1.37), but the 95% CI included 1 (0.99-1.89), compatible with no average association; IZ was 74.1% and the 95% prediction interval 0.59-3.17, so direction in a new setting is not predictable. High-risk-of-bias results carried 77.72% of the weight. Certainty was **very low** (Table 2).

**Figure 2.**
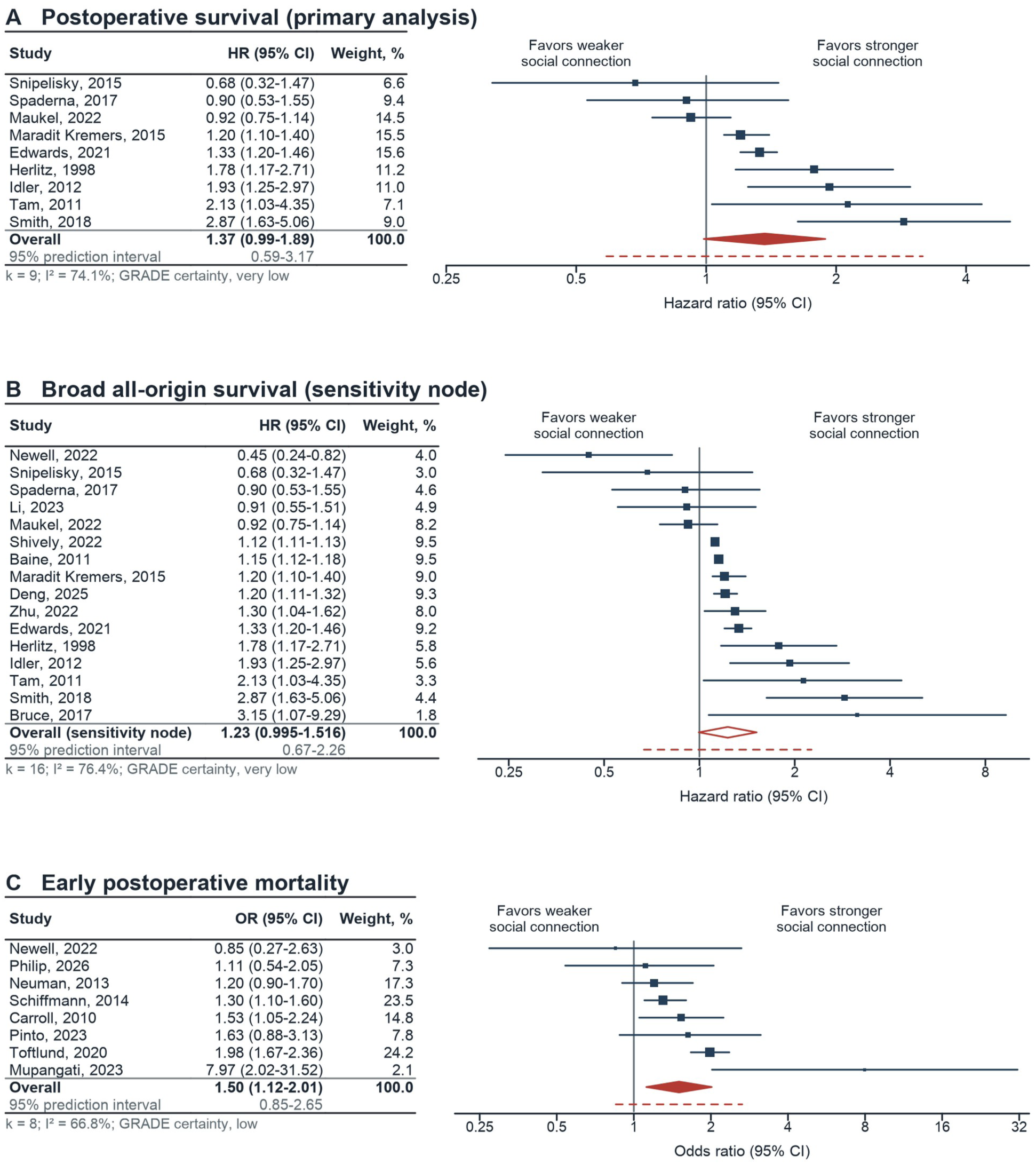
Preoperative Social Connection and Postoperative Survival and Early Mortality. Forest plots of A, postoperative all-cause survival, strict primary node (k = 9; HR, 1.37; 95% CI, 0.99-1.89; IZ, 74.1%; 95% prediction interval, 0.59-3.17; GRADE, very low); B, broad all-origin survival, maximum-coverage sensitivity node (k = 16; HR, 1.23; 95% CI, 0.995-1.516; IZ, 76.4%; 95% prediction interval, 0.67-2.26; GRADE, very low), which mixes follow-up time origins and is not an independent finding (open diamond; its CI is shown to 3 decimals, as in the text, because the lower bound rounds to 1.00); and C, early postoperative mortality (k = 8; OR, 1.50; 95% CI, 1.12-2.01; IZ, 66.8%; 95% prediction interval, 0.85-2.65; GRADE, low). Rows are labeled by first author and year. Squares indicate study estimates, with area proportional to random-effects weight within each panel (not comparable across panels; in A, weights are determined by the deposited standard errors and the node’s between-unit variance and reproduce the high-risk-of-bias share in Table 2). Horizontal lines indicate 95% CIs; each axis range contains every interval. Diamonds indicate pooled restricted maximum likelihood random-effects estimates with modified Hartung-Knapp 95% CIs (open diamond, sensitivity node), and dashed horizontal lines the 95% prediction intervals; the solid vertical line marks the null. Effects are oriented as weaker vs stronger preoperative social connection; values greater than 1 indicate worse outcomes with weaker connection (for survival, HR >1 = higher hazard of death), as the labels above each plot indicate. k denotes independent analysis weight units; k, IZ and GRADE certainty are printed beneath each table. CI indicates confidence interval; GRADE, Grading of Recommendations Assessment, Development and Evaluation; HR, hazard ratio; IZ, percentage of total variability attributable to between-study heterogeneity; OR, odds ratio.

### Broad all-origin survival (sensitivity analysis)

The maximum-coverage analysis (Figure 2B) included 16 units: HR, 1.23 (95% CI, 0.995-1.516; IZ = 76.4%; prediction interval 0.67-2.26), **very low** certainty; it mixes time origins, contains the strict node’s units and is not an independent finding.

### Early postoperative mortality

Across 8 analysis weight units (Figure 2C), the pooled association was OR, 1.50 (95% CI, 1.12-2.01; IZ = 66.8%; prediction interval 0.85-2.65, including the null). At baseline risks of 1% to 5%, the pooled OR corresponds to 5 to 23 additional deaths per 1000; explaining it away would require an unmeasured confounder associated with exposure and outcome at 2.29 to 2.35 each (CI bound, 1.47 to 1.48; eTable 7). High-risk-of-bias results carried 51.93% of the weight. Certainty was **low**.

### Non-home discharge

Across 9 analysis weight units (Figure 3A), weaker preoperative social connection was associated with higher odds of non-home discharge (OR, 1.95; 95% CI, 1.35-2.81; IZ = 78.1%). This was the largest association, but its prediction interval (0.71-5.34) was the widest and included 1. At baseline risks of 10% to 30%, the pooled OR corresponds to 78 to 155 additional non-home discharges per 1000 (E-values 2.40-2.96, CI-bound minimum 1.74; eTable 7). Discharge home depends partly on the resources defining the exposure. High-risk-of-bias results carried 50.79% of the weight. Certainty was **low**.

**Figure 3.**
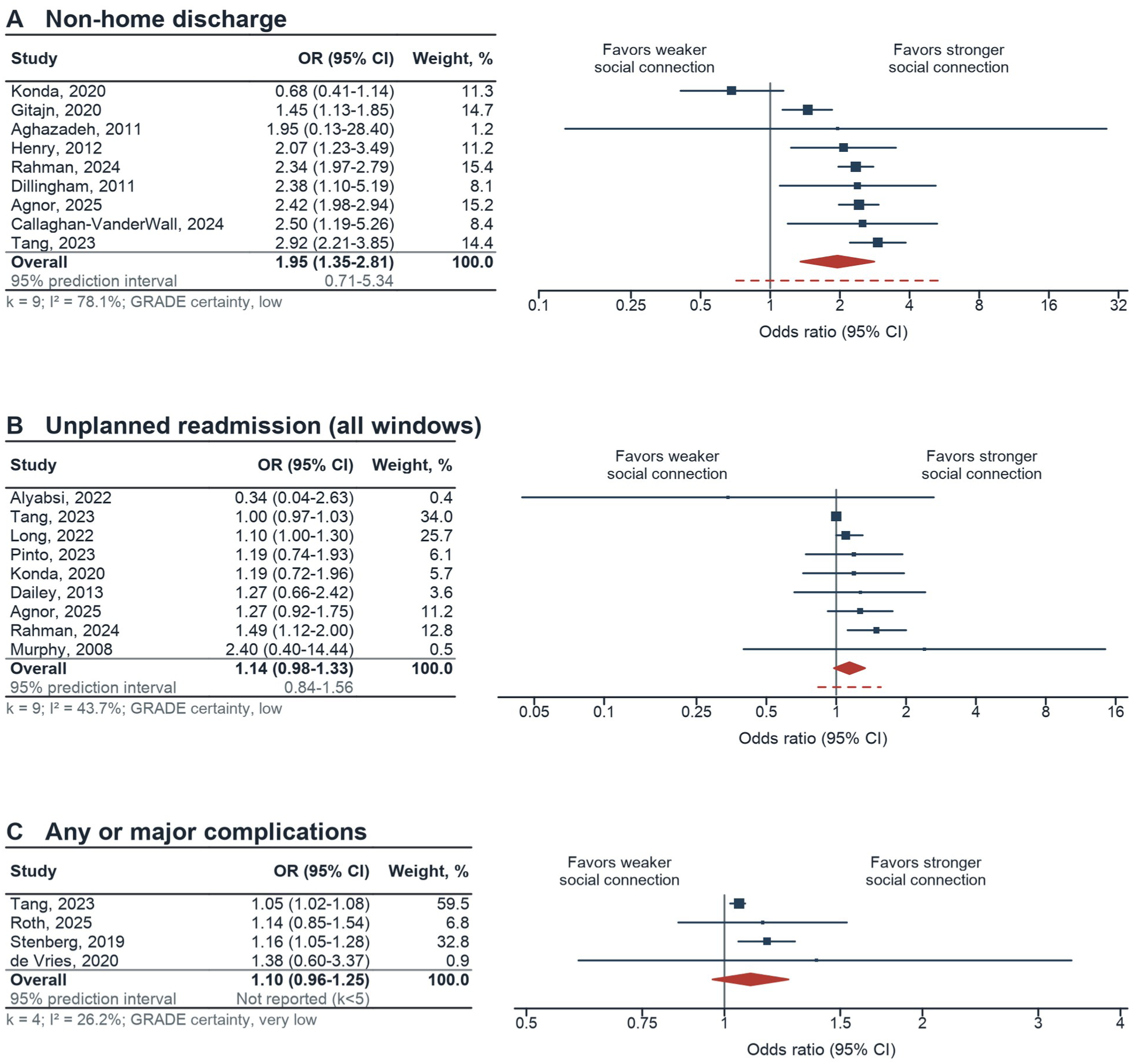
Preoperative Social Connection and Discharge Destination, Readmission, and Complications. Forest plots of A, non-home discharge (k = 9; OR, 1.95; 95% CI, 1.35-2.81; IZ, 78.1%; 95% prediction interval, 0.71-5.34; GRADE, low); B, unplanned readmission across all eligible windows (k = 9; OR, 1.14; 95% CI, 0.98-1.33; IZ, 43.7%; 95% prediction interval, 0.84-1.56; GRADE, low); and C, any or major postoperative complications (k = 4; OR, 1.10; 95% CI, 0.96-1.25; IZ, 26.2%; prediction interval not reported because k<5, a prespecified threshold; GRADE, very low). Orientation and drawing conventions: rows are labeled by first author and year; effects are oriented as weaker vs stronger preoperative social connection, with values greater than 1 indicating worse outcomes with weaker connection, as the labels above each plot indicate; k denotes independent analysis weight units and is printed with IZ and GRADE certainty beneath each table; squares are scaled to random-effects weight within each panel; horizontal lines indicate 95% CIs, and each axis range contains every interval; diamonds, pooled restricted maximum likelihood random-effects estimates with modified Hartung-Knapp 95% CIs; dashed horizontal lines, 95% prediction intervals (none in C because k<5); the solid vertical line, the null. CI indicates confidence interval; GRADE, Grading of Recommendations Assessment, Development and Evaluation; IZ, percentage of total variability attributable to between-study heterogeneity; OR, odds ratio.

### Unplanned readmission across all eligible windows

Across 9 analysis weight units (Figure 3B), the pooled point estimate was above 1 (OR, 1.14), but the 95% CI included 1 (0.98-1.33; IZ = 43.7%; prediction interval 0.84-1.56), **low** certainty: compatible with no average association. A stricter analysis of one uniformly defined readmission window could not be supported: under narrower eligibility (source-established unplanned status and time origin, an odds ratio, an eligible adjustment target and model, result-level risk of bias below high) no exact window was shared by more than 1 independent unit, below the 5 the protocol required. The strict-window question is therefore an evidence gap, not a null result.

### Complications

Across 4 analysis weight units of an any-or-major complication composite (Figure 3C), the pooled point estimate was above 1 (OR, 1.10) but the 95% CI included 1 (0.96-1.25; IZ = 26.2%; prediction interval not reported, k<5), compatible with no average association. All contributing results were at high risk of bias (100.0% of weight), and certainty was **very low**.

### Stability

No principal analysis changed direction on leave-one-out or under any legal alternative selection; none crossed the null for non-home discharge (0/15) or early mortality (0/18), whereas 71/72 crossed for readmission and 14/256 for the broad survival node (the locked selection among them), as did the sole complication selection (1/1; eTable 4). Restricting post hoc to marital or partnership contrasts left both CI-excluding associations essentially unchanged (OR, 1.48 and 1.87; eTable 3): in substance they are marital-status associations to which the umbrella contrast added 2 and 1 nonmarital units (fixed-effect and leave-one-out results, eTable 8).

### Risk of bias and certainty

Per-result judgments, eFigure 3; GRADE domains, eTable 9 and eTable 10. High-risk-of-bias results carried 50.79% to 100.0% of the random-effects weight; confounding and statistical reporting were the most compromised domains, and no analysis reached moderate certainty (Table 2). Exploratory funnel plots showed no asymmetry for the broad all-origin survival node (Egger P = .14) but asymmetry in a healthcare-burden family fitted only for dispersion diagnostics (k = 19; not an effect claim; Egger P = .005; eTable 11; eFigure 4); neither establishes nor excludes publication bias.

### Evidence gaps

Two gaps are structural: failure to rescue, where 8 reports analyzed the outcome and 0 estimated its association with preoperative social connection—an exposure–outcome cross gap, not an absent outcome (eTable 12)—and the strict-window readmission question.

## Discussion

In 445 studies, only 72 contributed a quantitative estimate to any fitted synthesis, and the primary survival analysis rested on 9 independent units. Point estimates were above 1 throughout, but the confidence interval excluded 1 only for early postoperative mortality and non-home discharge, certainty was low or very low, and every estimable prediction interval included the null. Those 2 associations are low-certainty findings rather than hypotheses: their direction survived leave-one-out, every legal alternative selection (0/18 and 0/15 crossed the null) and marital-status restriction (eTable 3). The literature supports a potential prognostic signal with substantial uncertainty, not a quantity ready for the bedside.

Three features explain that assessment.

### The unit of evidence is smaller than the unit of publication

The gap arises from shared registries, unanalyzed exposures and incompatible time origins (eMethods 2; eTable 2); counting analysis weight units rather than studies separates an apparently mature literature from a thin one. Seeing it required carrying the whole corpus through one eligibility contract, not a sample: every deduplicated record screened, every sought report accounted for, a source locator behind every analyzed estimate, risk of bias judged per principal-analysis result. Agent execution under investigator directives made that scope feasible; the meta-analysis itself is conventional.

### Constructs are not interchangeable

The umbrella families average over whatever each study measured, and marital status—the coarsest proxy for post-discharge resources—dominates. In general-population work, multidimensional measures carry larger mortality associations than binary marital status (OR, 1.91 vs 1.28)^1^, so construct choice plausibly attenuates these averages while residual confounding inflates them. Outcome definitions behave likewise: readmission windows differ.

### Heterogeneity materially limited transportability

For early postoperative mortality and non-home discharge the average association was positive, yet both prediction intervals (0.85-2.65 and 0.71-5.34), the range of true associations expected in new settings, crossed the null; each association may be absent in a future setting, which is why certainty is not higher.

This pattern is hypothesis-generating interpretation only: the largest association was for non-home discharge, the outcome most directly determined by whether a patient has somewhere to go, and the smallest for readmission and complications, driven largely by in-hospital factors operating before social resources matter. A mechanism acting after discharge would predict this, but the corpus does not test that mechanism, and socioeconomic confounding predicts it equally well.

General-population meta-analyses report similar magnitudes (OR, 1.50 here vs 1.50 for all-cause mortality^1^); concordance supports direction, not certainty. Cancer-cohort syntheses of marital status agree^57^, and single-specialty surgical reviews pool patient-reported outcomes or length of stay^58,59^; none addresses time origins, discharge destination, failure to rescue or registry dependence. The surgical social-determinants literature is far larger than the 72 studies usable here^60,61^, mostly measuring social connection without analyzing it.

### What would change these conclusions

Certainty would rise if studies prespecified social connection with a named instrument, defined outcome windows uniformly, reported time origins and the analyzed sample, and declared registry provenance—none of which requires a trial. Only failure to rescue needs new primary research: exposure and outcome were never estimated together in an eligible study.

For practice the defensible reading is narrow. Social connection is already recorded in many surgical cohorts, so the step is to read it, not collect it: where it is weak, the preoperative visit is the point to plan the discharge destination, name a caregiver and arrange follow-up; geriatric guidance already asks surgeons to assess support.^5^ These estimates do not support restricting access to surgery, steering patients from home discharge, or any risk-adjustment weight, and do not establish that changing social connection would change outcomes (not directly evaluated; Table 2).

## Limitations

First, all evidence is observational, and confounding was the most compromised QUIPS domain. Second, adjustment targets were mixed (Table 2); third, exposure ascertainment was dominated by binary marital status, usually incidental to the source study’s question. Fourth, asymmetry testing was possible in only 2 families, one of which showed asymmetry. Fifth, no overall participant total is reportable: sizes are additive only within a family, most are upper bounds and 1 estimate remains unsized. Sixth, screening and extraction were executed by AI agents under investigator directives; human confirmation of the 39 principal-analysis records was unblinded to AI-proposed values, so agreement cannot distinguish accuracy from anchoring, no record had independent duplicate human extraction, 18 sizes were checked, each by one investigator, and 27 (9 recovered bounds, 18 from extraction) were not; locators, replication and public data make this auditable. Seventh, no subgroup analysis or meta-regression was possible: no principal family reached k≥10. Eighth, 252 sought reports were not retrieved (Figure 1) and registered citation searching was not performed (eTable 1; eTable 13).

### Conclusions

Pooled point estimates were above 1 across all five principal outcomes, but confidence intervals excluded the null only for early postoperative mortality and non-home discharge—in this literature, predominantly marital-status associations—certainty was low or very low throughout, and every estimable prediction interval was compatible with no association in a new setting. The priority is not another meta-analysis of the same reports but primary studies that prespecify social connection as an exposure, define outcomes and time origins uniformly, and report failure to rescue.

## Supporting information

Supplementary Online Content

## Data Availability

The frozen analysis registries, membership tables, dependence-cluster registry, result-level risk-of-bias assessments, GRADE evidence profiles, sensitivity analyses, figure source data and deterministic analysis code underlying every reported number are publicly available now in the GitHub repository github.com/yinchuan123/social-connection-surgery-meta (code MIT license; data CC BY 4.0), archived on Zenodo with version DOI 10.5281/zenodo.22739144; no individual participant data were collected, and no copyrighted full-text articles are redistributed (eAppendix 1).

## Author Contributions

Dr Yin had full access to all of the data in the study and takes responsibility for the integrity of the data and the accuracy of the data analysis. Drs Yin and Wang contributed equally as co –first authors. *Concept and design:* Yin. *Acquisition, analysis, or interpretation of data:* Yin, Wang, Jiang, Jing. *Drafting of the manuscript:* Yin. *Critical review of the manuscript for important intellectual content:* All authors. *Statistical analysis:* Yin. *Administrative, technical, or material support:* Yin, Wang, Jiang, Huang. *Supervision:* Zhang, Jing.

## Artificial intelligence

Claude Code (Anthropic, PBC), a command-line large language model (LLM) agent running the models claude-fable-5, claude-opus-4-8, claude-sonnet-5, claude-opus-5 and claude-fable-5-1, was used between July 18 and September 13, 2026 as part of the formal research methods (Methods; eMethods 3; eAppendix 2) and to draft and revise the manuscript text, tables and supplementary materials from the frozen analytic release; LLM agents generated provisional structured assessments and source-grounded recommendations under frozen specifications, and the investigators adjudicated material ambiguities and authorized the final analytic release. The release number of the agent runtime was not retained. An OpenAI ChatGPT consumer subscription was used for general conceptual and strategic discussion only and did not generate or edit any content of this manuscript.

References were neither generated nor formatted by an LLM; reference metadata were retrieved from PubMed and Crossref records. Dr Yin personally purchased all AI subscriptions; no model provider had any role in the study. The authors reviewed all AI-generated content, verified every reported value against the signed analytic release, and take full responsibility for it. No AI system is an author.

## Additional Contributions

None.

## Prior Presentation

None.

