## Supplementary Online Content for "Preoperative Social Connection and Postoperative Outcomes in Adults Undergoing Surgery: A Systematic Review and Meta-analysis"

Registration: PROSPERO CRD420261449181. Analytic provenance identifier: SR-49523c19b885c87a (see eAppendix 1).

#### Contents


| Item | Title |
|---|---|
| eTable 1 | Registered protocol (PROSPERO CRD420261449181, Version 1.0, published July 14, 2026) versus conducted analysis |
| eMethods 1 | Complete search strategies and search amendment |
| eMethods 2 | Units of analysis, dependence handling and estimate selection |
| eMethods 2 addendum | Registry-provenance adjudication of SEER-derived clusters |
| eTable 2 | Contributing estimates of the principal analyses and of the broad all-origin survival node |
| eTable 3 | Post hoc marital-status-restricted sensitivity analyses |
| eMethods 3 | Artificial intelligence disclosure |
| eMethods 4 | Two-person human confirmation of source-grounded recommendations |
| eMethods 5 | Analyzed sample size: verification pass (September 2026) |
| eTable 4 | Selection-multiverse accounting and interval-method diagnostics by family |
| eTable 5 | PRISMA count reconciliation |
| eFigure 1 | Evidence landscape |
| eTable 6 | Characteristics of the 39 studies contributing to the principal analyses (33) or only to the broad all-origin survival sensitivity node (6) |
| eFigure 2 | Summary of the principal analyses |
| eTable 7 | Absolute risk differences and E-values (post hoc reader aids) |
| eTable 8 | Principal and prespecified sensitivity analyses |
| eFigure 3 | Result-level QUIPS risk of bias |
| eTable 9 | Result-level risk of bias (QUIPS) summary |
| eTable 10 | Formal prognostic certainty (GRADE, Claim B) by family and domain |
| eTable 11 | Small-study-effect diagnostics |
| eFigure 4 | Contour-enhanced funnel plots (k ≥ 10 families) |
| eTable 12 | Failure-to-rescue exposure-outcome cross-gap |
| eTable 13 | Clinical and review-process limitations |
| eAppendix 1 | Data and code availability |
| eAppendix 2 | Artificial intelligence: directive and prompt sequence |
| eReferences | References for studies cited in the Supplement |

### eTable 1. Registered protocol (PROSPERO CRD420261449181, Version 1.0, published July 14, 2026) versus conducted analysis

Every departure was decided blind to effect direction and dated before the synthesis lock (dated prespecification audit in the public release); registered wording is quoted from the public record. An administrative update was submitted to PROSPERO on September 8, 2026 (Version 2.0, published September 9, 2026), adding the collaborators who joined the review to the registered team; the funding source (none) and every eligibility, outcome, and analysis item were unchanged, and this table remains the itemized record of analytic departures.

| Item | Registered (Version 1.0) | Conducted | Direction and handling |
|---|---|---|---|
| Exposure pooling | "Structural (marital/partner status, living alone, objective isolation), perceived (loneliness), and functional (social support) exposure dimensions will never be pooled together." | Single umbrella contrast (weaker vs stronger connection) fixed in the frozen analysis plan before synthesis; each estimate carries its construct on a frozen axis (Table 1); nonmarital constructs contributed at most 2 units per family, and a post hoc marital-status-restricted sensitivity analysis is reported (eTable 3). | Documented departure; widens clinical heterogeneity and is priced into GRADE (indirectness/inconsistency); disclosed in Methods. |
| Primary outcome designation | Question triad: perioperative all-cause mortality (in-hospital, 30-day, 90-day), major complications (Clavien-Dindo grade >=III), and failure to rescue. | Primary: postoperative all-cause survival, strict time-origin node. Early mortality and complications are principal families; failure to rescue had no eligible exposure-outcome estimate (eTable 12). | Designation reassigned before synthesis lock, blind to results; all registered outcomes remain reported or accounted for; disclosed in Methods. |
| Risk-of-bias tool | Newcastle-Ottawa Scale. | Result-level QUIPS with formal prognostic GRADE. | Changed before assessment: the exposure is usually a covariate in the source studies, which the Newcastle-Ottawa Scale does not interrogate; QUIPS is the prognosis-specific instrument. |
| Interval estimation | DerSimonian-Laird intervals primary where few studies contribute; Hartung-Knapp-Sidik-Jonkman as conservative check. | REML with modified Hartung-Knapp (variance inflator floored at 1) primary in all families; DerSimonian-Laird and fixed-effect retained in the release. | Conservative direction (see eTable 4: where the floor bound, it widened intervals and removed one nominally positive finding). |
| Adjustment-based eligibility | Excludes "studies reporting only unadjusted estimates or lacking a measure of variance." | Adjustment carried as a frozen per-estimate axis; unadjusted estimates admitted where they survived the source-based hierarchy (composition in Table 2). | Documented departure; adjustment mix disclosed per family; variance requirement retained. |
| Dimension-specific secondary questions | Secondary questions for perceived and functional support dimensions. | Nonmarital constructs contributed at most 2 units per family; reported as evidence gaps rather than pooled. | Under-delivery disclosed (Limitations); no construct-specific claim is made. |
| Citation searching | Backward and forward citation searching of all included studies. | Not performed; identification relied on the 7 database searches alone (search reconciliation table in the public release). | Disclosed in Methods; a pre-execution validation against 15 known eligible studies (eMethods 1) partially served the registered sensitivity check. |
| Screening and data | "Studies will be screened | Title/abstract screening by | Machine involvement was |

| | | | |
|---|---|---|---|
| extraction | independently by at least two people (or person/machine combination) with a process to resolve differences"; records "screened in two stages (title/abstract, then full text) ... by two reviewers independently, with disagreements resolved by discussion or a third reviewer"; data "extracted independently by at least two people (or person/machine combination)". | 2 independent large-language-model agents blinded to each other (κ = 0.87), with a third agent adjudicating the 598 disagreements under investigator supervision; full-text assessment by agents with third-agent adjudication of disagreements, investigator rulings on protocol ambiguities and adversarial re-review of a risk-weighted sample of decisions; extraction by agents with source locators; two-person human confirmation of the 39 principal-analysis source records (unblinded to agent-proposed values); no independent human screening or extraction of the full pool. | anticipated by the registration ("person/machine combination"), but the registered two-reviewer process with third-reviewer resolution was implemented with agents rather than people; disclosed in Methods, eMethods 3 and Limitations; agent accuracy against a human reference standard was not measured. |
| Analyzed sample size (data item) | Data "extracted independently by at least two people (or person/machine combination) with a process to resolve differences"; no sample-size data item or tier convention is specified. | Not among the 8 human-confirmed fields; established after the synthesis lock (September 3-4, 2026) by one large-language-model agent with independent re-derivation by a second, in a fixed tier order (model N; arm sum; analyzed cohort; enclosing cohort) with upper bounds marked ≤; not human-confirmed (eMethods 5; eTable 2). | Post-lock descriptive addition; no membership, weight, estimate or interval depends on it; disclosed in Methods and Limitations. |
| Post hoc additions after the synthesis lock | Not in the registered record. | Marital-status-restricted sensitivity analyses (eTable 3; September 2, 2026) and reader aids (absolute risk differences and E-values, eTable 7), both derived from the locked inputs at the principal investigator's direction after internal peer-review-style audit. | Labelled post hoc wherever reported; no frozen number altered; no prediction interval reported for post hoc fits. |
| Review team and funding source (administrative) | Team: Dr Chuan Yin (guarantor and contact), Dr Zehao Jing; funding: "Review has no funding and no agreed support from an academic institution." | Six authors (title page); the study received no external funding and was funded personally by Dr Yin (Dr Jing is supported by National Natural Science Foundation of China grants 82302684 and 82572712 outside the submitted work). | Administrative update submitted to PROSPERO on September 8, 2026 (Version 2.0, published September 9, 2026); funding status consistent with the record (no study-specific funding); no analysis item affected. |

### eMethods 1. Complete search strategies and search amendment

Searches were executed on July 16-17, 2026 with no date or language limits. The strategy combined three blocks (social connection exposure; surgical/perioperative population; perioperative outcomes). One revision (revision D) was made before execution: validation against 15 known eligible studies showed that the draft strategy missed two of them, and the outcome and population blocks were therefore broadened (for example, functional-recovery outcome terms and fracture-cohort population terms); the final strategy retrieved all 15 validation items in PubMed. The revision preceded all screening; no database was searched after July 17, 2026.

#### PubMed/MEDLINE (NLM) — 4,813 records

```
("marital status"[Title/Abstract] OR "marital status"[MeSH Terms] OR "marriage status"[Title/Abstract]

 OR "unmarried"[Title/Abstract] OR "widowed"[Title/Abstract] OR "divorced"[Title/Abstract]

 OR "living alone"[Title/Abstract] OR "living arrangement"[Title/Abstract] OR "living arrangements"[Title/Abstract]

 OR "Social Isolation"[Title/Abstract] OR "Social Isolation"[MeSH Terms]

 OR "Loneliness"[Title/Abstract] OR "Loneliness"[MeSH Terms] OR "lonely"[Title/Abstract]

 OR "Social Support"[Title/Abstract] OR "Social Support"[MeSH Terms]

 OR "social network"[Title/Abstract] OR "cohabitation"[Title/Abstract]

 OR "unpartnered"[Title/Abstract] OR "partner status"[Title/Abstract] OR "spousal support"[Title/Abstract])

AND

("surg*"[Title/Abstract] OR "surgical procedures, operative"[MeSH Terms] OR "perioperative"[Title/Abstract]

 OR "postoperative"[Title/Abstract] OR "operative"[Title/Abstract] OR "arthroplasty"[Title/Abstract]

 OR "resection"[Title/Abstract]

 OR "hip fracture"[Title/Abstract] OR "hip fractures"[MeSH Terms] OR "fracture"[Title/Abstract]

 OR "Perioperative Period"[MeSH Terms] OR "Postoperative Period"[MeSH Terms])

AND

("mortality"[Title/Abstract] OR "death"[Title/Abstract] OR "failure to rescue"[Title/Abstract]

 OR "complication*"[Title/Abstract] OR "readmission"[Title/Abstract] OR "length of stay"[Title/Abstract]

 OR "patient discharge"[MeSH Terms] OR "discharge disposition"[Title/Abstract]

 OR "non-home discharge"[Title/Abstract] OR "morbidity"[Title/Abstract] OR "Postoperative Complications"[MeSH Terms]

 OR "delirium"[Title/Abstract] OR "Delirium"[MeSH Terms]

 OR "functional recovery"[Title/Abstract] OR "Recovery of Function"[MeSH Terms]

 OR "activities of daily living"[Title/Abstract] OR "Activities of Daily Living"[MeSH Terms]

 OR "patient-reported outcome*"[Title/Abstract] OR "care home"[Title/Abstract]

 OR "nursing home"[Title/Abstract] OR "discharge destination"[Title/Abstract])
```

#### Embase (Elsevier; Emtree) — 15,002 records

```
('marital status'/exp OR 'social isolation'/exp OR 'loneliness'/exp OR 'social support'/exp OR 'social network'/exp OR 'cohabitation'/exp OR 'single person'/exp
```

OR 'marital status':ti,ab OR 'marriage status':ti,ab OR unmarried:ti,ab OR widowed:ti,ab OR divorced:ti,ab

OR (living NEAR/1 alone):ti,ab OR 'living arrangement':ti,ab OR 'living arrangements':ti,ab

OR 'social isolation':ti,ab OR loneliness:ti,ab OR lonely:ti,ab OR 'social support':ti,ab OR 'social network':ti,ab

OR cohabitation:ti,ab OR unpartnered:ti,ab OR 'partner status':ti,ab OR 'spousal support':ti,ab)

AND

('surgery'/exp OR 'surgical technique'/exp OR 'perioperative period'/exp OR 'postoperative period'/exp

OR 'hip fracture'/exp OR 'fracture'/exp

OR surg*:ti,ab OR perioperative:ti,ab OR postoperative:ti,ab OR operative:ti,ab OR arthroplasty:ti,ab OR resection:ti,ab

OR 'hip fracture':ti,ab OR fracture:ti,ab)

AND

('mortality'/exp OR 'postoperative complication'/exp OR 'hospital readmission'/exp OR 'length of stay'/exp OR 'hospital discharge'/exp OR 'failure to rescue'/exp

OR 'delirium'/exp OR 'convalescence'/exp OR 'daily life activity'/exp OR 'nursing home'/exp OR 'patient-reported outcome'/exp

OR mortality:ti,ab OR death:ti,ab OR 'failure to rescue':ti,ab OR complication*:ti,ab OR readmission:ti,ab

OR 'length of stay':ti,ab OR 'discharge disposition':ti,ab OR 'non-home discharge':ti,ab OR morbidity:ti,ab

OR delirium:ti,ab OR 'functional recovery':ti,ab OR 'activities of daily living':ti,ab

OR 'patient reported outcome*':ti,ab OR 'care home':ti,ab OR 'nursing home':ti,ab OR 'discharge destination':ti,ab)

#### CINAHL Complete (EBSCOhost) — 2,739 records

( (MH "Marital Status+") OR (MH "Social Isolation") OR (MH "Loneliness") OR (MH "Support, Psychosocial")

OR (MH "Social Networks") OR (MH "Living Arrangements") OR (MH "Family+")

OR TI("marital status" OR "marriage status" OR unmarried OR widowed OR divorced OR "living alone" OR "living arrangement*" OR "social isolation" OR loneliness OR lonely OR "social support" OR "social network*" OR cohabitation OR unpartnered OR "spousal support")

OR AB("marital status" OR "marriage status" OR unmarried OR widowed OR divorced OR "living alone" OR "living arrangement*" OR "social isolation" OR loneliness OR lonely OR "social support" OR "social network*" OR cohabitation OR unpartnered OR "spousal support") )

AND

( (MH "Surgery, Operative+") OR (MH "Perioperative Care+") OR (MH "Postoperative Period") OR (MH "Hip Fractures") OR (MH "Fractures, Bone+")

OR TI(surg* OR perioperative OR postoperative OR operative OR arthroplasty OR resection OR "hip fracture*" OR fracture)

OR AB(surg* OR perioperative OR postoperative OR operative OR arthroplasty OR resection OR "hip fracture*" OR fracture) )

AND

( (MH "Postoperative Complications+") OR (MH "Mortality+") OR (MH "Readmission") OR (MH "Length of Stay") OR (MH "Patient Discharge+")

OR (MH "Delirium") OR (MH "Activities of Daily Living+") OR (MH "Nursing Homes")

OR TI(mortality OR death OR "failure to rescue" OR complication* OR readmission OR "length of stay" OR "discharge disposition" OR morbidity OR delirium OR "functional recovery" OR "activities of daily living" OR "patient-reported outcome*" OR "care home" OR "nursing home" OR "discharge destination")

OR AB(mortality OR death OR "failure to rescue" OR complication* OR readmission OR "length of stay" OR "discharge disposition" OR morbidity OR delirium OR "functional recovery" OR "activities of daily living" OR "patient-reported outcome*" OR "care home" OR "nursing home" OR "discharge destination") )

#### APA PsycInfo (EBSCOhost) — 634 records

( DE "Marital Status" OR DE "Social Isolation" OR DE "Loneliness" OR DE "Social Support" OR DE "Social Networks"

OR DE "Cohabitation" OR DE "Marriage" OR DE "Widows" OR DE "Divorced Persons"

OR TI("marital status" OR "marriage status" OR unmarried OR widowed OR divorced OR "living alone" OR "living arrangement*" OR "social isolation" OR loneliness OR lonely OR "social support" OR "social network*" OR cohabitation OR unpartnered OR "spousal support")

OR AB("marital status" OR "marriage status" OR unmarried OR widowed OR divorced OR "living alone" OR "living arrangement*" OR "social isolation" OR loneliness OR lonely OR "social support" OR "social network*" OR cohabitation OR unpartnered OR "spousal support") )

AND

( DE "Surgery" OR DE "Postsurgical Complications"

OR TI(surg* OR perioperative OR postoperative OR operative OR arthroplasty OR resection OR "hip fracture*" OR fracture)

OR AB(surg* OR perioperative OR postoperative OR operative OR arthroplasty OR resection OR "hip fracture*" OR fracture) )

AND

( DE "Delirium" OR DE "Activities of Daily Living"

OR TI(mortality OR death OR "failure to rescue" OR complication* OR readmission OR "length of stay" OR "discharge disposition" OR morbidity OR recovery OR delirium OR "activities of daily living" OR "patient-reported outcome*" OR "care home" OR "nursing home")

OR AB(mortality OR death OR "failure to rescue" OR complication* OR readmission OR "length of stay" OR "discharge disposition" OR morbidity OR recovery OR delirium OR "activities of daily living" OR "patient-reported outcome*" OR "care home" OR "nursing home") )

#### Web of Science Core Collection (Clarivate; TS=Topic) — 3,076 records

TS=("marital status" OR "marriage status" OR unmarried OR widowed OR divorced OR "living alone" OR "living arrangement*" OR "social isolation" OR loneliness OR lonely OR "social support" OR "social network*" OR cohabitation OR unpartnered OR "partner status" OR "spousal support")

AND

TS=(surg* OR perioperative OR postoperative OR operative OR arthroplasty OR resection OR "hip fracture*" OR fracture*)

AND

TS=(mortality OR death OR "failure to rescue" OR complication* OR readmission OR "length of stay" OR "discharge disposition" OR "non-home discharge" OR morbidity OR delirium OR "functional recovery" OR "activities of daily living" OR "patient-reported outcome*" OR "care home" OR "nursing home" OR "discharge destination")

#### Scopus (Elsevier; TITLE-ABS-KEY) — 6,485 records

TITLE-ABS-KEY("marital status" OR "marriage status" OR unmarried OR widowed OR divorced OR "living alone" OR "living arrangement*" OR "social isolation" OR loneliness OR lonely OR "social support" OR "social network*" OR cohabitation OR unpartnered OR "partner status" OR "spousal support")

AND

TITLE-ABS-KEY(surg* OR perioperative OR postoperative OR operative OR arthroplasty OR resection OR "hip fracture*" OR fracture*)

AND

TITLE-ABS-KEY(mortality OR death OR "failure to rescue" OR complication* OR readmission OR "length of stay" OR "discharge disposition" OR "non-home discharge" OR morbidity OR delirium OR "functional recovery" OR "activities of daily living" OR "patient-reported outcome*" OR "care home" OR "nursing home" OR "discharge destination")

#### CENTRAL (Cochrane Library; Search Manager) — 552 records

([mh "Marital Status"] OR [mh "Social Isolation"] OR [mh Loneliness] OR [mh "Social Support"] OR [mh "Social Networking"]

OR ("marital status" OR "marriage status" OR unmarried OR widowed OR divorced OR "living alone" OR (living NEXT arrangement*) OR "social isolation" OR loneliness OR lonely OR "social support" OR (social NEXT network*) OR cohabitation OR unpartnered):ti,ab,kw)

AND

([mh "Surgical Procedures, Operative"] OR [mh "Perioperative Period"] OR [mh "Hip Fractures"]

OR (surg* OR perioperative OR postoperative OR operative OR arthroplasty OR resection OR (hip NEXT fracture*) OR fracture*):ti,ab,kw)

AND

([mh "Postoperative Complications"] OR [mh "Patient Readmission"] OR [mh "Length of Stay"] OR [mh "Patient Discharge"]

OR [mh Delirium] OR [mh "Recovery of Function"] OR [mh "Activities of Daily Living"]

OR (mortality OR death OR "failure to rescue" OR complication* OR readmission OR "length of stay" OR "discharge disposition" OR morbidity OR delirium OR (functional NEXT recovery) OR "activities of daily living" OR (patient NEXT reported NEXT outcome*) OR "care home" OR "nursing home"):ti,ab,kw)

### eMethods 2. Units of analysis, dependence handling and estimate selection

Reports (publications), studies, cohort entities (source-defined cohorts after collapsing duplicate reporting), dependence clusters (cohort entities known or conservatively judged to share participants) and analysis weight units (the independent units receiving weight in a model) were tracked separately; k denotes analysis weight units throughout. Within a dependence cluster, one weight unit entered a given analysis. Where a study reported more than one eligible estimate for an outcome, a single estimate entered each analysis through a frozen source-based selection hierarchy that never used effect direction, interval width or whether an estimate crossed the null; every legal alternative selection was re-run as a multiverse analysis (eTable 4). Ratio estimates were analyzed on the natural-log scale with standard errors derived from the printed confidence limits, and estimates indexed on the stronger-connection group were inverted so that every contrast reads as weaker versus stronger connection, as the registered protocol required; the per-estimate comparator adjudication is deposited. Across the included corpus these quantities are 445 studies, 441 cohort entities and 421 dependence clusters.

Outcomes were assigned to a frozen controlled vocabulary before synthesis; the definitions governing the principal analyses are reproduced verbatim below. Time origin and follow-up window are carried as separate per-estimate axes (eTable 2), so an outcome label never fixes a horizon.

| Code | Definition |
|---|---|
| All cause mortality | Death from any cause within the specified window measured from the specified time origin. |
| Unscheduled readmission | Rehospitalisation not planned at index discharge. |
| Non home discharge | Discharge to any destination other than the private home. |
| Major complication | Composite of complications meeting a stated severity threshold. |
| Any complication | Composite of any postoperative complication irrespective of severity. |

Independent software reproduction. All principal random-effects models were refitted in R 4.6.1 (metafor 5.0.1 and meta 8.5.0) from the deposited per-unit log effects and standard errors. Pooled estimates, 95% CIs (restricted maximum likelihood with the modified Hartung-Knapp adjustment) and $I^2$ values matched the reported values to the reported precision in both packages; prediction intervals matched when computed with the registered formula (t distribution with k-2 degrees of freedom and the modified standard error), whereas the meta package default, which uses an unmodified standard error, differs in the second decimal for the 2 families in which the modification applied. The script and the row-by-row record accompany the data deposit.

### eMethods 2 addendum. Registry-provenance adjudication of SEER-derived clusters

Four dependence clusters rest on cohorts derived from the Surveillance, Epidemiology, and End Results (SEER) program. Their per-cluster adjudications (verbatim, with evidence fragments) are deposited in the public independent-cohort adjudication registry; in summary: one cluster pair was judged SAME_COHORT (two reports of the same analysis, values identical digit-for-digit) and collapsed to one unit; three clusters were judged PARTIAL_OVERLAP on same-registry, same-disease, intersecting-window evidence without a reported shared-participant count, and were conservatively collapsed rather than treated as independent. Misjudged clusters can only merge units, deflating k and widening intervals.

### eTable 2. Contributing estimates of the principal analyses and of the broad all-origin survival node

One row per analysis weight unit: 39 estimate rows (from 33 studies) of the five principal families, all drawn from the human-confirmed membership lock, followed by the 7 units that enter only the broad all-origin survival sensitivity node. Exposure contrasts and follow-up windows are verbatim from the printed sources. The construct column classifies the analyzed contrast, not merely what a study measured; the frozen hierarchy has no loneliness category, so the single single-item loneliness contrast (Herlitz) was left unclassified rather than mapped post hoc (eFigure 1B counts it under network/isolation as measured). The strict survival node fixes the time origin, not the horizon. Analyzed sizes follow a fixed tier order, shown per row in Size basis: the size printed for the fitted model (Model N); the summed sizes of the two arms of the contrast where both are printed (Arm sum); the analyzed cohort after stated exclusions (Analyzed cohort); or the enclosing cohort where the source shows attrition it never counts. A size is exact only when it is the fitted model's own printed denominator, or an arm sum from the estimate's own table whose arms exhaust the analyzed set with no further exclusion stated; every other size, including all sizes from the original extraction pass (cohort totals verified against their locator only), is an upper bound marked ≤. The same printed count can therefore be exact for one estimate and a bound for another. A dagger marks sizes established in the September 2026 passes; of these, only the first-pass sizes were human-signed and no analyzed size, in either pass, was confirmed by the two-person human pass (eMethods 5). A double dagger marks units whose count is procedures or discharges rather than unique patients. Pinto reports different denominators for in-hospital mortality (879) and 30-day readmission (890) within one 896-patient cohort, as printed. Sizes may be summed within a family, whose units are non-overlapping dependence clusters, but never across families, because a cohort can contribute to more than one family. Units whose time origin or window the source did not establish are retained in the all-windows readmission family and are ineligible for any exact-window analysis.

| Family | Study | Exposure contrast (verbatim) | Construct | Adjustment | Time origin | Follow-up window | Analyzed N | Size basis | Overall RoB |
|---|---|---|---|---|---|---|---|---|---|
| Early postoperative mortality | Carroll | Not Married (marital status) | Marital/ partnership | Post-exposure-conditioned | Index operation | 30 days from resection | ≤1,847 | Enclosing cohort (original pass) | High |
| Early postoperative mortality | Philip | Not lonely, socially isolated (baseline four-group classification) | Network/ isolation | Post-exposure-conditioned | Index operation | 90 days; time origin = index surgery date | ≤27,905† | Analyzed cohort (September 2026 pass) | Moderate |
| Early postoperative mortality | Newell | Married (marital status) | Marital/ partnership | Baseline-adjusted | Hospital admission | In-hospital or 30 days | ≤785† | Analyzed cohort (September 2026 pass) | High |
| Early postoperative mortality | Neuman | Widowed (marital status, SEER, recorded at diagnosis) | Marital/ partnership | Post-exposure-conditioned | Index operation | 90 days from surgery | ≤12,979 | Enclosing cohort (original pass) | Moderate |
| Early postoperativ | Schiffmann | Unmarried marital status | Marital/ partnership | Baseline-adjusted | Index operation | 90 days (from surgery) | 5,207† | Model N (Septemb | Moderate |

| | | | | | | | | | |
|---|---|---|---|---|---|---|---|---|---|
| e mortality | | | | | | | | er 2026 pass) | |
| Early postoperative mortality | Pinto | Estado civil = Viúvo (widowed) | Marital/ partnership | Unadjusted | Hospital admission | In-hospital (index stay + any 30-day readmission stay); time origin = hospital admission for surgery | ≤879 | Enclosing cohort (original pass) | High |
| Early postoperative mortality | Toftlund | Marital status: Unmarried | Marital/ partnership | Baseline-adjusted | Index operation | 30 days | ≤13,795† | Analyzed cohort (September 2026 pass) | High |
| Early postoperative mortality | Mupangati | Residence status: living alone | Living arrangement | Post-exposure-conditioned | Hospital admission | In-hospital / index elective-surgery admission; no fixed day-count | ≤382 | Enclosing cohort (original pass) | High |
| Complications | Stenberg | Marital status: Divorced/widow/ widower | Marital/ partnership | Unadjusted | Index operation | 30 days from surgery | ≤40,156† | Analyzed cohort (September 2026 pass) | High |
| Complications | Roth | Living alone (n=212) | Living arrangement | Unadjusted | Index operation | Postoperative, min 3 months follow-up | ≤1,466† | Analyzed cohort (September 2026 pass) | High |
| Complications | de Vries | Marital status: Widow | Marital/ partnership | Unadjusted | Index operation | within 30 days after surgery | ≤151† | Enclosing cohort (September 2026 pass) | High |
| Complications | Tang | Married marital status | Marital/ partnership | Mixed/unclear | Index operation | 90 days postoperative | ≤106,752† | Analyzed cohort (September 2026 pass) | High |
| Non-home discharge | Dillingham | Married | Marital/ partnership | Post-exposure-conditioned | Hospital admission | Immediate post-acute disposition (admission within 3 days of acute-care discharge), ascertained over the 6-month post- | ≤348† | Analyzed cohort (September 2026 pass) | Moderate |

| | | | | | | | | | |
|---|---|---|---|---|---|---|---|---|---|
| | | | | | | index-amputation reference period | | | |
| Non-home discharge | Konda | Marital status = Divorced | Marital/ partnership | Post-exposure-conditioned | Hospital admission | Disposition at end of index hospitalisation | ≤1,931 | Enclosing cohort (original pass) | High |
| Non-home discharge | Henry | Unmarried marital status (single, separated, divorced, or widowed), ascertained preoperatively | Marital/ partnership | Post-exposure-conditioned | Ambiguous (multiple anchors reported) | At discharge from the index valve-surgery admission | ≤307 | Enclosing cohort (original pass) | Moderate |
| Non-home discharge | Tang | Married marital status | Marital/ partnership | Mixed/unclear | Not established in source | at index discharge | ≤106,752 † | Analyzed cohort (September 2026 pass) | High |
| Non-home discharge | Aghazadeh | Single/ widowed/ divorced (unmarried) marital status | Marital/ partnership | Post-exposure-conditioned | Hospital admission | Discharge status at end of index hospitalization after radical cystectomy | ≤440† | Analyzed cohort (September 2026 pass) | High |
| Non-home discharge | Rahman | Marital status: Divorced | Marital/ partnership | Baseline-adjusted | Not established in source | At discharge | ≤14,462† ‡ | Analyzed cohort (September 2026 pass) | High |
| Non-home discharge | Agnor | Living alone | Living arrangement | Baseline-adjusted | Hospital admission | At discharge from index surgical hospitalization | ≤3,248 | Enclosing cohort (original pass) | Moderate |
| Non-home discharge | Callaghan-VanderWall | Marriage status: Single/divorced/ widowed | Marital/ partnership | Unadjusted | Hospital admission | Discharge disposition of the index surgical admission; time origin = surgery/index admission | ≤135 | Enclosing cohort (original pass) | High |
| Non-home discharge | Gitajn | Marital status: unmarried (divorced, widowed, separated) | Marital/ partnership | Mixed/unclear | Hospital admission | At discharge from the initial hospitalization (time origin = index admission) | ≤2,365 | Enclosing cohort (original pass) | Moderate |
| Postoperativ | Smith | Lower | Perceived/ | Unadjusted | Equivalent | Median follow- | ≤273† | Enclosing | High |

| | | | | | | | | | |
|---|---|---|---|---|---|---|---|---|---|
| e survival (strict node) | | Perceived Social Support (PSSS), median split, with transplant LOS >=1 month | functional support | | index event (device implantation or transplantation) | up 6.1 years (range 0-13.4) from transplant | | cohort (September 2026 pass) | |
| Postoperative survival (strict node) | Maukel | Limited social support (clinician-judged, applicable) | Perceived/ functional support | Post-exposure-conditioned | Equivalent index event (device implantation or transplantation) | Full follow-up from CF-LVAD implantation (median 15.1 mo); time-averaged HR | 8,471† | Model N (September 2026 pass) | High |
| Postoperative survival (strict node) | Tam | Married at time of OHT listing (social-worker-recorded marital status; stable-partner cohabitants counted as married) | Marital/ partnership | Baseline-adjusted | Equivalent index event (device implantation or transplantation) | 5 years from transplant, conditional on surviving 60 days | ≤260† | Enclosing cohort (September 2026 pass) | High |
| Postoperative survival (strict node) | Edwards | Living alone (cohabitation status) | Living arrangement | Baseline-adjusted | Index operation | 365 days post-THA | 96,764† | Arm sum (September 2026 pass) | Moderate |
| Postoperative survival (strict node) | Spaderna | Unmarried (marital status at time of listing) | Marital/ partnership | Unadjusted | Equivalent index event (device implantation or transplantation) | Post-HTx, median follow-up 70 months (range <1-93 months) | 148† | Model N (September 2026 pass) | High |
| Postoperative survival (strict node) | Herlitz | NHP item 9 "I feel lonely" = yes | Other/ unclassified | Mixed/unclear | Index operation | 5 years after CABG (time origin: surgery) | ≤1,270† | Arm sum (September 2026 pass) | High |
| Postoperative survival (strict node) | Idler | Not married (aggregated) | Marital/ partnership | Unadjusted | Index operation | >=3 months after surgery to end of follow-up; 87 deaths | 545† | Model N (September 2026 pass) | High |
| Postoperative survival (strict node) | Maradit Kremers | Marital status: Divorced/widowed (2809, 14%) | Marital/ partnership | Baseline-adjusted | Index operation | Any time during follow-up, time origin = surgery (procedures 2002-2009; registry follow-up >65% complete at 30 years) | ≤20,124 | Enclosing cohort (original pass) | High |
| Postoperative survival (strict node) | Snipelisky | Married/living with a partner (marital status, | Marital/ partnership | Baseline-adjusted | Equivalent index event (device | Time-to-death from LVAD implantation | ≤136 | Enclosing cohort (original | Moderate |

| | | | | | | | | | |
|---|---|---|---|---|---|---|---|---|---|
| | | preoperative) | | | implantation or transplantation) | through Sept 30, 2014 (mean follow-up 2.2 years) | | pass) | |
| Unplanned readmission | Dailey | Marital status = Divorced/separated | Marital/ partnership | Unadjusted | Hospital discharge | 30 days from discharge | ≤3,261†‡ | Model N (September 2026 pass) | High |
| Unplanned readmission | Konda | Marital status = Divorced | Marital/ partnership | Post-exposure-conditioned | Not established in source | 90 days (time origin not stated) | ≤1,931 | Enclosing cohort (original pass) | High |
| Unplanned readmission | Long | Marital status: Single | Marital/ partnership | Baseline-adjusted | Hospital discharge | 30 days (Methods clock: after discharge) | ≤167,265 †‡ | Analyzed cohort (September 2026 pass) | Moderate |
| Unplanned readmission | Alyabsi | Marital status = Married | Marital/ partnership | Unadjusted | Hospital discharge | 30 days from index discharge | ≤356 | Enclosing cohort (original pass) | High |
| Unplanned readmission | Tang | Married marital status | Marital/ partnership | Mixed/unclear | Not established in source | not specified | ≤106,752 † | Analyzed cohort (September 2026 pass) | High |
| Unplanned readmission | Rahman | Marital status: Divorced | Marital/ partnership | Baseline-adjusted | Index operation | 90 days after surgery | ≤14,462† ‡ | Analyzed cohort (September 2026 pass) | High |
| Unplanned readmission | Pinto | Estado civil = Viúvo (widowed) | Marital/ partnership | Unadjusted | Hospital discharge | 30 days after hospital discharge (time origin = discharge, not surgery/admission) | ≤890 | Enclosing cohort (original pass) | High |
| Unplanned readmission | Murphy | Living alone | Living arrangement | Baseline-adjusted | Hospital discharge | 30 days from hospital discharge | 175† | Model N (September 2026 pass) | High |
| Unplanned readmission | Agnor | Living alone | Living arrangement | Baseline-adjusted | Not established in source | 30 days (origin not stated) | 3,248† | Model N (September 2026 pass) | Moderate |
| Broad all-origin survival node only | Bruce | Patient lives alone | Living arrangement | Baseline-adjusted | Not fixed (all-origin node) | Time-to-event from hospital discharge, censored at | ≤61 | Enclosing cohort (original pass) | High |

| | | | | | | | | | |
|---|---|---|---|---|---|---|---|---|---|
| (sensitivity) | | | | | | transplant or 2015-12-31 (up to ~6 y) | | | |
| Broad all-origin survival node only (sensitivity) | Shively | Marital status: Not Married | Marital/ partnership | Unadjusted | Not fixed (all-origin node) | 5 years from diagnosis | ≤120,598 † | Enclosing cohort (September 2026 pass) | High |
| Broad all-origin survival node only (sensitivity) | Li | Marital status: single/divorced/ widowed | Marital/ partnership | Unadjusted | Not fixed (all-origin node) | To 36-month endpoint; median follow-up 36 months (range 3.3-36); 77 deaths in the full cohort of 320 | ≤320 | Enclosing cohort (original pass) | High |
| Broad all-origin survival node only (sensitivity) | Newell | Married women (marital status, female stratum) | Marital/ partnership | Mixed/unclear | Not fixed (all-origin node) | Presented as the adjusted counterpart of the 1-year Kaplan-Meier analysis; the Methods state no time restriction for the Cox model and median total follow-up was 2.4 years — the exact horizon is NOT established in the text | Not reported | Not printed | High |
| Broad all-origin survival node only (sensitivity) | Zhu | Widowed marital status | Marital/ partnership | Post-exposure-conditioned | Not fixed (all-origin node) | Latest follow-up (median 37.1 months, from admission) | ≤1,101† | Analyzed cohort (September 2026 pass) | Moderate |
| Broad all-origin survival node only (sensitivity) | Baine | Married at time of diagnosis | Marital/ partnership | Post-exposure-conditioned | Not fixed (all-origin node) | 3 years from diagnosis | ≤34,555 | Enclosing cohort (original pass) | Moderate |
| Broad all-origin survival node only (sensitivity) | Deng | Marital status = Married | Marital/ partnership | Unadjusted | Not fixed (all-origin node) | Whole follow-up (max 212 months); time origin not defined in text | ≤15,109 | Enclosing cohort (original pass) | High |

### eTable 3. Post hoc marital-status-restricted sensitivity analyses

Authorized post hoc by the principal investigator (September 2, 2026), after peer-review-style internal audit. Each row refits the family restricted to units whose analyzed contrast is marital or partnership status (classification shown per unit in eTable 2), using the identical frozen per-estimate effects and the identical REML plus modified Hartung-Knapp machinery; the full six locked analyses were first refitted from the same frozen inputs and matched the locked values exactly (positive control). No frozen number was altered. Both confidence-interval-excluding umbrella associations were materially unchanged under the restriction. The broad all-origin node is omitted here as it is not an independent finding; prediction intervals are not reported for these post hoc fits.

| Family | Umbrella k | Umbrella estimate (95% CI) | Marital-restricted k | Marital-restricted estimate (95% CI) | Marital-restricted $I^2$, % |
|---|---|---|---|---|---|
| Postoperative survival (strict node) | 9 | 1.37 (0.99-1.89) | 5 | 1.27 (0.75-2.17) | 60.4 |
| Early postoperative mortality | 8 | 1.50 (1.12-2.01) | 6 | 1.48 (1.12-1.96) | 66.0 |
| Non-home discharge | 9 | 1.95 (1.35-2.81) | 8 | 1.87 (1.21-2.90) | 79.6 |
| Unplanned readmission | 9 | 1.14 (0.98-1.33) | 7 | 1.12 (0.94-1.33) | 47.3 |
| Complications | 4 | 1.10 (0.96-1.25) | 3 | 1.09 (0.89-1.34) | 47.6 |

### eMethods 3. Artificial intelligence disclosure

#### A. Artificial intelligence in the research methods

Large-language-model (LLM) agents (Claude Code; Anthropic, PBC) were used between July 18 and September 13, 2026 as part of the formal research methods. Recorded model identifiers were claude-fable-5, claude-opus-4-8, claude-sonnet-5, claude-opus-5 and claude-fable-5-1; the release number of the agent runtime was not retained and is disclosed as a reproducibility limitation; reproduction of every reported result depends on the frozen ledgers, source locators and deterministic analysis code in the deposit, not on re-running the agents. LLM agents generated provisional structured assessments and source-grounded recommendations under frozen specifications; investigators adjudicated material ambiguities and authorized the final analytic release. Agent tasks comprised record management, title and abstract screening (2 independent agents blinded to each other, with disagreements adjudicated by a third agent), full-text eligibility assessment with adversarial re-review, full-text retrieval bookkeeping, structured extraction of estimates into controlled ledgers, construction of the exposure ontology, outcome definitions and dependence-cluster registry, drafting of the deterministic analysis code, and generation of the source-grounded recommendations that were then confirmed by two people. Specifications were versioned as numbered written investigator directives together with hash-frozen protocols (eligibility criteria, prognostic GRADE protocol; the readmission protocol is archived with the project); major prompt sequences followed the project phases (screening, extraction, dependence adjudication, synthesis, audit, release), and outputs were revised only through further numbered directives. Error mitigation included frozen controlled vocabularies, release gates that must pass before promotion, adversarial re-review of findings (a separate verifier agent, holding the full text, re-judged a risk-weighted sample of full-text decisions and locked findings, seeking grounds to overturn them; disagreements between screening agents were adjudicated by a third agent, protocol ambiguities by investigator rulings followed by agent re-review, and residual cases by the investigators), independent statistical replication of all locked results, and verification of every analyzed estimate against a structured locator to the printed source. The frozen registries, code, and the eligibility and GRADE protocols are contained in the reproducibility deposit package accompanying the repository record. Record management, the controlled ledgers, source-locator verification, release gates and the generation of source-grounded recommendations were implemented in Evidence OS, evidence-management software developed and owned by Dr Yin (see Conflict of Interest Disclosures); the deterministic analysis and build code deposited with the signed release is the software state used. The numbered investigator directives and agent prompt templates are retained in the project archive and are available from the corresponding authors on request; their sequence, dates, governing frozen specifications and revision mechanism are set out in eAppendix 2. They are not part of the public deposit because they quote the copyrighted full texts they operated on; the deposit therefore carries the frozen protocols, registries, source locators and deterministic code, which are what reproduction of the reported results requires. The accuracy of agent screening decisions against a human reference standard was not measured: the reported κ is agreement between two agents, and the third agent that adjudicated disagreements saw both agents' decisions.

Copyrighted content entered into the model. Bibliographic records exported from the licensed databases and the full texts of the reports assessed for eligibility were entered into the agent runtime for screening and extraction. Full texts were obtained through the authors' institutional library subscriptions, under open-access licenses, or by individual purchase through paid document-delivery services, and were used under the respective access terms for the internal purposes of eligibility assessment and data extraction only; no publisher-specific permission for machine processing was sought beyond those terms. No copyrighted full text is reproduced in the manuscript, the supplement or the public deposit, which carries no article full text: only extracted data elements with structured locators to the printed sources, together with the frozen protocols, registries and deterministic code (eAppendix 1).

#### B. Artificial intelligence in manuscript preparation

The same tools were used to draft and revise the manuscript text, tables and supplementary materials from the frozen analytic release; eFigures 1 to 4 were rendered by deterministic Python plotting code and Figures 2 and 3 by deterministic R code (forestploter), all from the frozen release and the locked values; the PRISMA flow diagram (Figure 1) is a static vector graphic drawn from the frozen counts, and no generative image tool was used. Bibliographic references were neither generated nor formatted by an LLM; all reference metadata were retrieved from PubMed and Crossref records. The authors reviewed all AI-generated content, verified every reported value against the signed analytic release, and take full responsibility for the content of the manuscript. No AI system is listed as an author. Under the provider's terms of service, rights in generated output rest with the user, and the authors hold the rights to publish the AI-assisted text.

| Item | Value | Status |
|---|---|---|
| Platform / tool | Claude Code (command-line LLM agent) | Recorded |
| Manufacturer | Anthropic, PBC | Recorded |
| Recorded model identifiers | claude-fable-5; claude-opus-4-8; claude-sonnet-5; claude-opus-5; claude-fable-5-1 | Recorded from execution logs |
| Exact runtime build identifier | Not retained; disclosed as a reproducibility limitation | Not retained (disclosed as a limitation) |
| Dates of use | 2026-07-18 to 2026-09-13 | Recorded from execution logs |

| | | |
|---|---|---|
| Risk-of-bias and certainty assessment | Not recorded as an agent task. The two study-level risk-of-bias assessors are identified in the deposit only by reviewer identifiers, so the record does not establish whether they were investigators or agents; result-level judgments were derived from the study level by frozen rule and certainty was computed from the frozen GRADE protocol | Recorded (identifiers only) |
| Second vendor (not part of the research methods) | OpenAI ChatGPT consumer subscription: general conceptual and strategic discussion only; generated or edited no manuscript content and performed no screening, extraction, assessment or analysis | Recorded (scope only; no execution log) |
| Specification versioning | Numbered investigator directives + hash-frozen protocols | Recorded |
| Human adjudication | Investigators adjudicated material ambiguities via numbered written directives and authorized the final analytic release; two-person confirmation of all 39 principal-family source records | Recorded |
| Source verification | Structured source locators for every analyzed estimate; deterministic locator verification; independent statistical replication of locked results | Recorded |
| Copyrighted content entered into the model | Yes: licensed bibliographic records and the full texts of reports assessed for eligibility, obtained through institutional library subscriptions, open-access licenses or paid document-delivery purchase, and used under the respective access terms for screening and extraction only; no publisher-specific machine-processing permission sought; no full text redistributed | Disclosed (Methods; eMethods 3) |
| Rights in AI-generated content | Output rights rest with the user under the provider terms of service; the authors hold the rights to the AI-assisted text | Disclosed |
| AI listed as an author | No | Confirmed |

### eMethods 4. Two-person human confirmation of source-grounded recommendations

All 39 source records underlying the principal analyses underwent a two-person human confirmation of source-grounded recommendations. For each record, the Evidence OS pipeline generated one source-grounded recommendation from the printed source, its structured locator, the frozen canonical ledgers and the family definitions, together with any supported alternative, a confidence statement and the expected scientific impact. Recommendation logic did not use effect direction, statistical significance, whether an interval crossed the null, whether a study strengthened or weakened the pooled result, or whether the resulting certainty grade would be more favorable.

Both reviewers worked in separate consoles with separated storage and exports, answered the same 8 field-level questions per record (source identity; exposure and comparator; outcome and timing; numerical effect; transformation; adjustment and model selection; cohort and dependence; analysis membership) and gave a final decision without seeing each other's answers. Both reviewers, however, saw the same Evidence OS recommendations; their agreement therefore does not demonstrate convergence of two independent judgment paths, and the process was not blinded independent dual review.

Results: both reviewers completed 39 of 39 records; agreement was 39/39 on the final decision and 312/312 on the field-level questions; there were no disagreements and no item required adjudication; no scientific correction of any kind resulted (no change to membership, k, pooled results, grades, dependence clusters or denominators).

### eMethods 5. Analyzed sample size: verification pass (September 2026)

The original source-verification pass confirmed eight field-level items per analyzed estimate; analyzed sample size was not among them, and the sizes it recorded were cohort totals verified against their locator only. In September 2026, after internal peer-review-style audit, the estimates without a size were revisited in a dedicated pass. For each, one large-language-model agent located the sample size of the model that produced that estimate in the printed full text and recorded a verbatim quotation with line numbers; a second, independent agent re-derived the size from the same source without adopting the first answer, and separately confirmed that the quotation appears at the stated lines. The two agreed on all 28 estimates examined. Sizes were established for 18 (specific to the fitted model in 7, the analyzed cohort after stated exclusions in 11); for 10, the printed source reports no size for that model: in 7 the model deleted or excluded cases the source never counts, in 2 the two arms are printed but never summed, and in 1 the estimate is a within-stratum contrast whose stratum total is never printed. A second pass then revisited those 10 estimates, recovering a bounding size in a fixed tier order: the summed sizes of the two contrast arms where both are printed (2); the analyzed cohort after stated exclusions (3); failing those, the enclosing cohort, which bounds the fitted size from above (4). Each recovered size was independently re-derived and its quotations re-checked line by line; 9 were recovered and 1 remains unsized (Newell, a within-female contrast whose stratum total the source never prints). A size is exact only when it is the fitted model's own printed denominator or an arm sum whose arms exhaust the analyzed set with no further exclusion stated; all other sizes are marked ≤, and family totals inherit that mark. Sizes established in either September pass carry a dagger in eTable 2; the full record, including both agents' answers and the quotations, is in the public archive (eAppendix 1).

Human sign-off. On September 5, 2026 the first-pass rows were checked against the printed sources by investigators working from a signing register that presented, for each estimate, the verbatim quotation, its file and line numbers, and the command that prints those lines. Each row was assigned to one investigator and signed by that person: 18 sizes and 10 findings that no model size was printed were signed by 2 investigators, with 0 corrections; a row without a signature was not counted as signed. The check was made by one investigator per row, not in duplicate, and the reviewers saw the proposed value while checking it. The 9 bounding sizes recovered in the second pass for those estimates were not presented for signature and remain agent-verified only; the remaining 18 sizes come from the original extraction pass, were not part of this sign-off, and remain cohort totals verified against their locator only. The signed register is in the public archive (eAppendix 1).

### eTable 4. Selection-multiverse accounting and interval-method diagnostics by family

A legal alternative selection substitutes, for any contributing cohort, another estimate tied under the frozen source-based hierarchy; the family denominator is the product of tie multiplicities, and every combination was evaluated. Null-crossing is judged on the modified Hartung-Knapp random-effects interval. The unmodified Hartung-Knapp interval is shown as a diagnostic: where the variance-inflator floor bound, it only widened intervals (for unplanned readmission the unmodified interval excluded the null while the reported floored interval did not), so the modification erred against, not for, the reported associations. The strict-node primary survival analysis has a unique frozen selection and therefore no selection multiverse; its stability rests on leave-one-out (0/9 direction flips). Three thresholds are distinct: the k≥2 fitting floor for any pooled model, the k≥5 threshold below which prediction intervals are not reported, and the supportability minimum of 5 independent units required before a strict-window readmission claim could be reported.

| Family | k | Candidate estimates | Legal selections | Selections crossing the null | Unmodified HK interval (diagnostic only) |
|---|---|---|---|---|---|
| Postoperative all-cause survival (primary, strict node) | 9 | unique frozen selection | not applicable | not applicable | not exported for this node |
| Broad all-origin survival (sensitivity node) | 16 | 70 | 256 | 14/256 | 0.9954-1.5160 |
| Early postoperative mortality | 8 | 40 | 18 | 0/18 | 1.1189-2.0138 |
| Non-home discharge | 9 | 22 | 15 | 0/15 | 1.3765-2.7638 |
| Unplanned readmission across all eligible windows | 9 | 27 | 72 | 71/72 | 1.0001-1.3011 |
| Any/major postoperative complications | 4 | 4 | 1 | 1/1 | 0.9972-1.2034 |
| Healthcare-burden dispersion diagnostics (not an effect claim) | 19 | 65 | 14976 | 0/14976 | 1.2151-1.8085 |

### eTable 5. PRISMA count reconciliation

| Stage | Count |
|---|---|
| Records identified (7 databases) | 33301 |
| Duplicates removed | 12902 |
| Records screened | 20399 |
| Excluded at title/abstract | 18420 |
| Reports sought for retrieval | 1979 |
| Reports not retrieved | 252 |
| Retrieved but not eligible for full assessment | 177 |

| | |
|---|---|
| Reports assessed for eligibility | 1550 |
| Excluded with reasons | 970 |
| Not included: eligibility not determinable from retrievable evidence | 102 |
| Not classifiable under the frozen protocol (terminal protocol-ambiguity states) | 24 |
| Reports of included studies | 454 |
| Studies included | 445 |

Exclusion reasons: Exposure not analyzed — Only in baseline table / covariate without own effect (n = 215); Ineligible exposure — Exposure not measured preoperatively (n = 200); Review/editorial/non-original report — Review / editorial / protocol / qualitative / case report (n = 159); Ineligible population — Non-adult / non-surgical / inseparable surgical subgroup (n = 155); Ineligible outcome — No outcome on the protocol list (n = 137); No eligible effect estimate — No extractable or derivable effect estimate (n = 44); Ineligible follow-up time — Only outcomes beyond 1 year (n = 30); Ineligible exposure — No social connection variable — only SES / insurance / race (n = 23); Ineligible exposure — Social support delivered as an intervention (n = 6); Review/editorial/non-original report — Qualitative / non-quantitative design (n = 1). "Retrieved but not eligible for full assessment" denotes reports whose retrieved files did not meet the locked requirements for entering full assessment (per-record dispositions are in the public release).

### eFigure 1. Evidence landscape

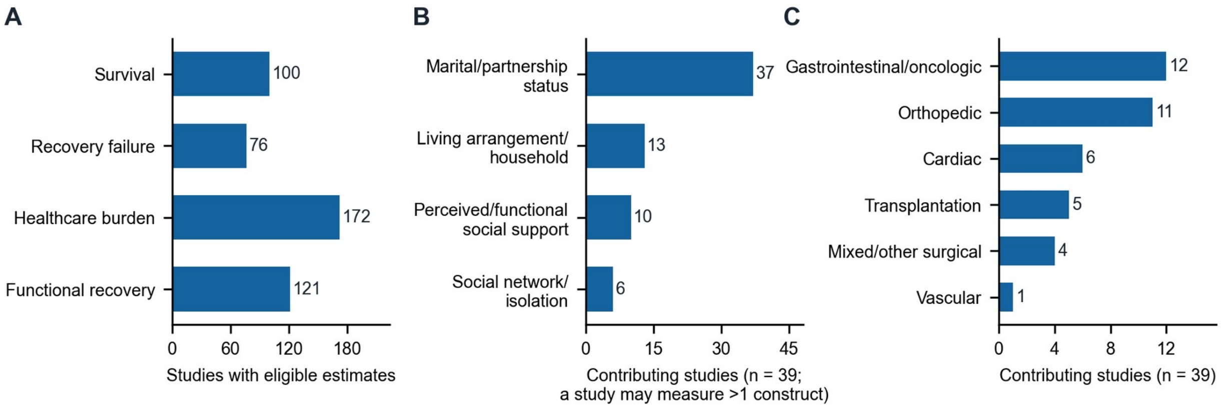


eFigure 1. Evidence landscape of preoperative social connection and postoperative outcomes. A, Studies with at least one extracted eligible estimate in each outcome domain, counted across all 445 included studies; a study may contribute to several domains, and domain counts must not be summed or read as subsets of any smaller set. B, Social connection constructs measured across the principal and sensitivity analyses (one study may measure more than one construct). C, Surgical specialty of studies contributing to the principal and sensitivity analyses. Panel A counts unique studies with eligible estimates across all 445 included studies; panels B and C count the 39 unique studies contributing to the principal and sensitivity analyses. Universes differ by design and must not be cross-tabulated. Panel A follows the prespecified outcome-domain order; panels B and C are sorted by count.

### eTable 6. Characteristics of the 39 studies contributing to the principal analyses (33) or only to the broad all-origin survival sensitivity node (6)

| Study ID | eRef | Year | Journal | Surgical specialty | Social connection construct | Contributes to | PMID |
|---|---|---|---|---|---|---|---|
| S00039 | e3 | 2018 | Transpl Int | Transplantation | Perceived/functional social support | Postoperative survival; Broad all-origin survival | 29130541 |
| S00320 | e14 | 2011 | J Heart Lung Transplant | Transplantation | Marital/partnership status; Living arrangement/household; Social network/isolation | Postoperative survival; Broad all-origin survival | 21907593 |
| S00384 | e21 | 2021 | Acta Orthop | Orthopaedic | Marital/partnership status; Living arrangement/household | Postoperative survival; Broad all-origin survival | 34085592 |
| S00410 | e23 | 2017 | J Am Heart Assoc | Transplantation | Marital/partnership status; Living arrangement/household; Perceived/functional social support; Social network/isolation | Postoperative survival; Broad all-origin survival | 29187384 |
| S00706 | e33 | 1998 | Eur J Vasc Endovasc Surg | Cardiac | Marital/partnership status; Living arrangement/household; Social network/isolation | Postoperative survival; Broad all-origin survival | 9728430 |
| S00751 | e34 | 2012 | J Health Soc Behav | Cardiac | Marital/partnership status; Perceived/functional social support; Social network/isolation | Postoperative survival; Broad all-origin survival | 22382719 |
| S00784 | e35 | 2015 | J Arthroplasty | Orthopaedic | Marital/partnership status | Postoperative survival; Broad all-origin survival | 25971779 |
| S01320 | e36 | 2015 | American Heart Journal | Transplantation | Marital/partnership status; Perceived/functional social support | Postoperative survival; Broad all-origin survival | 26542496 |
| S00269 | e10 | 2022 | J Am Heart Assoc | Cardiac | Marital/partnership status; Perceived/functional social support | Postoperative survival; Broad all-origin survival | 35191318 |
| S00053 | e4 | 2017 | Circ Cardiovasc Qual Outcomes | Transplantation | Marital/partnership status; Living arrangement/household; Perceived/functional social support; Social network/isolation | Broad all-origin survival | 28073849 |
| S00168 | e7 | 2022 | Cureus | Gastrointestinal/oncologic | Marital/partnership status | Broad all-origin survival | 35295347 |
| S00431 | e24 | 2011 | PLoS One | Gastrointestinal/oncologic | Marital/partnership status | Broad all-origin survival | 21698253 |
| S00332 | e15 | 2023 | Ir J Med Sci | Gastrointestinal/oncologic | Marital/partnership status | Broad all-origin survival | 36862310 |
| S00352 | e16 | 2022 | Am J Cardiol | Cardiac | Marital/partnership status; Living arrangement/household; Perceived/functional social support | Broad all-origin survival; Early postoperative mortality | 35361479 |
| S00393 | e22 | 2022 | Int J Gen Med | Orthopaedic | Marital/partnership status | Broad all-origin survival | 36238540 |
| S01462 | e38 | 2025 | Current Problems in Surgery | Gastrointestinal/oncologic | Marital/partnership status | Broad all-origin survival | 40306865 |
| S00237 | e8 | 2010 | J Gastrointest Surg | Gastrointestinal/oncologic | Marital/partnership status | Early postoperative mortality | 20844977 |
| S00245 | e9 | 2026 | Br J Anaesth | Mixed/other surgical | Living arrangement/household; | Early postoperative | 41136321 |

| | | | | | | | |
|---|---|---|---|---|---|---|---|
| | | | | | Perceived/functional social support; Social network/isolation | mortality | |
| S00355 | e17 | 2013 | Ann Surg Oncol | Mixed/other surgical | Marital/partnership status | Early postoperative mortality | 23292483 |
| S00535 | e28 | 2023 | Rev Bras Ortop (Sao Paulo) | Orthopaedic | Marital/partnership status | Early postoperative mortality; Unplanned readmission | 37252296 |
| S00567 | e29 | 2020 | Scand J Public Health | Mixed/other surgical | Marital/partnership status | Early postoperative mortality | 31973622 |
| S01620 | e39 | 2023 | Bali Medical Journal | Mixed/other surgical | Marital/partnership status; Living arrangement/household | Early postoperative mortality | |
| S00484 | e26 | 2014 | Eur J Surg Oncol | Gastrointestinal/oncologic | Marital/partnership status | Early postoperative mortality | 25454826 |
| S00011 | e1 | 2011 | PM R | Vascular | Marital/partnership status; Living arrangement/household; Perceived/functional social support | Non-home discharge | 21497320 |
| S00289 | e11 | 2012 | Ann Thorac Surg | Cardiac | Marital/partnership status | Non-home discharge | 22835556 |
| S00382 | e20 | 2023 | Front Oncol | Gastrointestinal/oncologic | Marital/partnership status | Non-home discharge; Unplanned readmission; Any/major complications | 37849800 |
| S00475 | e25 | 2011 | J Urol | Gastrointestinal/oncologic | Marital/partnership status; Living arrangement/household | Non-home discharge | 21074199 |
| S00692 | e32 | 2024 | Am J Phys Med Rehabil | Orthopaedic | Marital/partnership status | Non-home discharge | 38206613 |
| S00670 | e31 | 2025 | PLoS One | Orthopaedic | Marital/partnership status; Living arrangement/household | Non-home discharge; Unplanned readmission | 39746111 |
| S01378 | e37 | 2020 | Journal of Orthopaedic Trauma | Orthopaedic | Marital/partnership status | Non-home discharge | 31868766 |
| S00498 | e27 | 2024 | J Am Acad Orthop Surg | Orthopaedic | Marital/partnership status | Non-home discharge; Unplanned readmission | 38100772 |
| S00144 | e5 | 2020 | Geriatr Orthop Surg Rehabil | Orthopaedic | Marital/partnership status | Non-home discharge; Unplanned readmission | 32030312 |
| S00033 | e2 | 2013 | J Bone Joint Surg Am | Orthopaedic | Marital/partnership status | Unplanned readmission | 23780539 |
| S00309 | e12 | 2022 | Int J Surg | Orthopaedic | Marital/partnership status | Unplanned readmission | 35811014 |
| S00370 | e18 | 2022 | BMC Gastroenterol | Gastrointestinal/oncologic | Marital/partnership status | Unplanned readmission | 36229783 |
| S00596 | e30 | 2008 | Eur J Cardiovasc Prev Rehabil | Cardiac | Marital/partnership status; Living arrangement/household | Unplanned readmission | 18391650 |
| S00166 | e6 | 2019 | Surg Obes Relat Dis | Gastrointestinal/oncologic | Marital/partnership status | Any/major complications | 30826242 |
| S00313 | e13 | 2025 | J Plast Reconstr Aesthet Surg | Gastrointestinal/oncologic | Marital/partnership status; Living arrangement/household; Perceived/functional social support | Any/major complications | 40440986 |
| S00374 | e19 | 2020 | Eur J Surg Oncol | Gastrointestinal/oncologic | Marital/partnership status | Any/major complications | 31427138 |

### eFigure 2. Summary of the principal analyses

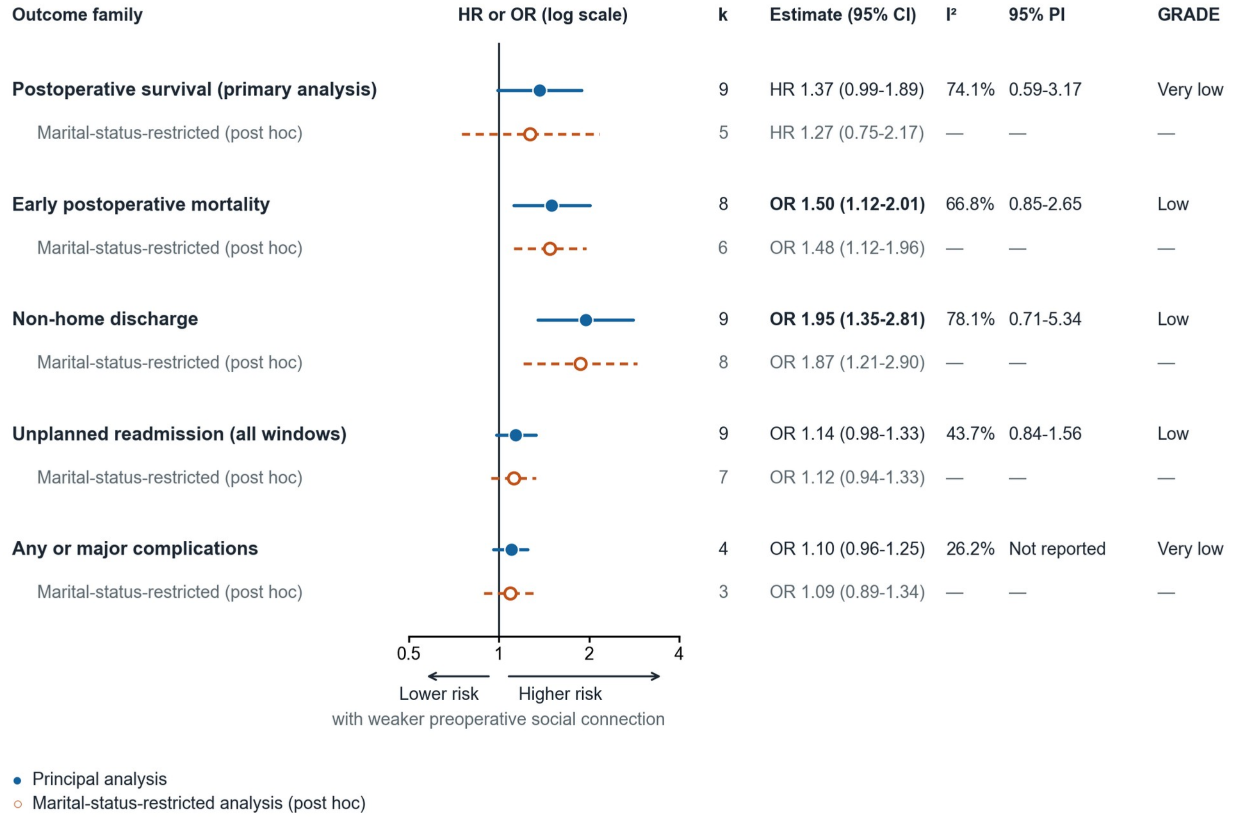


**eFigure 2. Summary of the principal analyses and the marital-status-restricted sensitivity analyses.** Of 445 mapped studies, 72 contributed poolable estimates; the principal analyses rest on 4 to 9 independent analysis weight units (k). Filled markers indicate the principal analyses (weaker vs stronger preoperative social connection, whichever construct a study measured; restricted maximum likelihood random effects with modified Hartung-Knapp 95% CIs); open markers indicate post hoc analyses restricted to units whose analyzed contrast was marital or partnership status (eTable 3). Values greater than 1 indicate worse outcomes with weaker connection; the solid vertical line marks the null. Bold text marks principal-analysis 95% CIs that exclude 1. $I^2$, 95% prediction intervals (reported where k≥5), and GRADE certainty are shown for the principal analyses only and apply to the umbrella prognostic-association claim (Claim B; eTable 10); —, not reported for the post hoc analyses. The broad all-origin survival node

and the 39 contributing studies are described in Figure 2B and eFigure 1. CI indicates confidence interval; GRADE, Grading of Recommendations Assessment, Development and Evaluation; HR, hazard ratio; I², percentage of total variability attributable to between-study heterogeneity; OR, odds ratio; PI, prediction interval.

### eTable 7. Absolute risk differences and E-values (post hoc reader aids)

| Analysis | Quantity | BaselineRisk | Value | Basis |
|---|---|---|---|---|
| Early postoperative mortality | E-value (point) | rare-outcome assumption (<5%) | 2.37 | OR 1.50 treated as RR; E = RR + sqrt(RR(RR-1)) |
| Early postoperative mortality | E-value (CI bound) | rare-outcome assumption (<5%) | 1.48 | CI lower limit 1.12 |
| Early postoperative mortality | E-value (point) | 1% | 2.35 | exact odds-to-risk conversion: RR 1.49 at baseline 1% |
| Early postoperative mortality | E-value (CI bound) | 1% | 1.48 | CI lower limit converts to RR 1.12 |
| Early postoperative mortality | E-value (point) | 3% | 2.32 | exact odds-to-risk conversion: RR 1.48 at baseline 3% |
| Early postoperative mortality | E-value (CI bound) | 3% | 1.47 | CI lower limit converts to RR 1.11 |
| Early postoperative mortality | E-value (point) | 5% | 2.29 | exact odds-to-risk conversion: RR 1.46 at baseline 5% |
| Early postoperative mortality | E-value (CI bound) | 5% | 1.47 | CI lower limit converts to RR 1.11 |
| Early postoperative mortality | Absolute risk difference per 1000 | 1% | +5 (95% CI, +1 to +10) | exact odds-to-risk conversion of locked OR and CI |
| Early postoperative mortality | Absolute risk difference per 1000 | 3% | +14 (95% CI, +3 to +29) | exact odds-to-risk conversion of locked OR and CI |
| Early postoperative mortality | Absolute risk difference per 1000 | 5% | +23 (95% CI, +6 to +46) | exact odds-to-risk conversion of locked OR and CI |
| Non-home discharge | E-value (point) | 10% | 2.96 | exact odds-to-risk conversion: RR 1.78 at baseline 10% |
| Non-home discharge | E-value (CI bound) | 10% | 1.94 | CI lower limit converts to RR 1.31 |
| Non-home discharge | Absolute risk difference per 1000 | 10% | +78 (95% CI, +31 to +138) | exact odds-to-risk conversion of locked OR and CI |
| Non-home discharge | E-value (point) | 20% | 2.66 | exact odds-to-risk conversion: RR 1.64 at baseline 20% |
| Non-home discharge | E-value (CI bound) | 20% | 1.84 | CI lower limit converts to RR 1.26 |
| Non-home discharge | Absolute risk difference per 1000 | 20% | +128 (95% CI, +53 to +213) | exact odds-to-risk conversion of locked OR and CI |
| Non-home discharge | E-value (point) | 30% | 2.40 | exact odds-to-risk conversion: RR 1.52 at baseline 30% |
| Non-home discharge | E-value (CI bound) | 30% | 1.74 | CI lower limit converts to RR 1.22 |
| Non-home discharge | Absolute risk difference per 1000 | 30% | +155 (95% CI, +67 to +247) | exact odds-to-risk conversion of locked OR and CI |

Derived post hoc, at the principal investigator's direction (September 2, 2026), from the locked pooled estimates only; no membership, weight or interval was recomputed. Odds-to-risk conversion is exact at each stated baseline risk (RR = OR/(1-p0+p0*OR)); the square-root approximation was not used. For early postoperative mortality both the rare-outcome value (OR treated as RR) and exact conversions at baseline risks of 1%, 3% and 5% are shown, because the baselines shown span the range the protocol treats as rare, and one contributing unit reports a 30-day mortality above that range;

for non-home discharge, where baseline risk is not rare, all quantities are shown across representative baselines. E-value = RR + sqrt(RR(RR-1)) (VanderWeele and Ding). All reader aids are computed from unrounded locked values.

### eTable 8. Principal and prespecified sensitivity analyses

| Analysis | Type | k | RE estimate | RE 95% CI | FE estimate | $I^2$ | 95% prediction interval | Leave-one-out direction flips | Selections crossing the null |
|---|---|---|---|---|---|---|---|---|---|
| Postoperative all-cause survival (primary) | Primary | 9 | 1.3653 | 0.9874-1.8879 | 1.2640 | 74.1% | 0.5874-3.1737 | 0/9 | Not applicable (unique frozen selection) |
| Broad all-origin survival (maximum-coverage sensitivity) | Sensitivity | 16 | 1.2284 | 0.9954-1.5160 | 1.1265 | 76.4% | 0.6680-2.2590 | 0/16 | 14/256 |
| Early postoperative mortality | Key secondary | 8 | 1.5011 | 1.1189-2.0138 | 1.5560 | 66.8% | 0.8503-2.6500 | 0/8 | 0/18 |
| Non-home discharge | Key secondary | 9 | 1.9504 | 1.3516-2.8146 | 2.1328 | 78.1% | 0.7128-5.3370 | 0/9 | 0/15 |
| Unplanned readmission across all eligible windows | Key secondary | 9 | 1.1407 | 0.9782-1.3302 | 1.0123 | 43.7% | 0.8360-1.5564 | 0/9 | 71/72 |
| Any/major postoperative complications | Key secondary | 4 | 1.0955 | 0.9594-1.2509 | 1.0602 | 26.2% | Not reported (k<5) | 0/4 | 1/1 |

k denotes independent analysis weight units. RE, random-effects (REML, modified Hartung-Knapp); FE, fixed-effect. The complete registry of all fitted local analyses is provided in the repository technical appendix accompanying the data deposit.

### eFigure 3. Result-level QUIPS risk of bias

| | Participation | Attrition | Exposure | Outcome | Confounding | Statistics | Overall |
|---|---|---|---|---|---|---|---|
| **Postoperative / broad survival** | | | | | | | |
| Baine, 2011 | M | M | M | L | M | M | M |
| Bruce, 2017 | H | H | H | M | H | H | H |
| Deng, 2025 | M | M | M | M | H | M | H |
| Edwards, 2021 | L | L | M | L | M | L | M |
| Herlitz, 1998 | H | H | M | M | M | H | H |
| Idler, 2012 | H | M | L | L | H | M | H |
| Li, 2023 | M | M | M | M | H | M | H |
| Maradit Kremers, 2015 | M | H | M | M | L | M | H |
| Maukel, 2022 | L | M | H | M | M | H | H |
| Newell, 2022 | L | M | M | L | M | H | H |
| Shively, 2022 | M | M | M | M | H | M | H |
| Smith, 2018 | H | ? | L | L | H | H | H |
| Snipelisky, 2015 | M | M | M | M | M | M | M |
| Spaderna, 2017 | M | L | L | L | H | M | H |
| Tam, 2011 | M | M | M | M | M | H | H |
| Zhu, 2022 | M | M | M | M | M | M | M |
| **Early postoperative mortality** | | | | | | | |
| Carroll, 2010 | M | M | M | L | M | H | H |
| Mupangati, 2023 | H | ? | M | M | M | H | H |
| Neuman, 2013 | M | ? | M | L | M | M | M |
| Newell, 2022 | L | M | M | L | M | H | H |
| Philip, 2026 | M | M | L | L | L | L | M |
| Pinto, 2023 | M | ? | M | M | H | M | H |
| Schiffmann, 2014 | M | M | M | L | M | M | M |
| Toftlund, 2020 | L | L | M | L | H | M | H |

| | Participation | Attrition | Exposure | Outcome | Confounding | Statistics | Overall |
|---|---|---|---|---|---|---|---|
| **Non-home discharge** | | | | | | | |
| Aghazadeh, 2011 | M | L | M | M | M | H | H |
| Agnor, 2025 | L | M | M | L | L | M | M |
| Callaghan-VanderWall, 2024 | M | ? | M | M | H | M | H |
| Dillingham, 2011 | L | M | M | M | M | M | M |
| Gitajn, 2020 | M | M | M | L | L | M | M |
| Henry, 2012 | M | M | M | M | M | M | M |
| Konda, 2020 | H | M | M | M | M | M | H |
| Rahman, 2024 | M | ? | M | M | M | H | H |
| Tang, 2023 | L | L | M | M | M | H | H |
| **Unplanned readmission** | | | | | | | |
| Agnor, 2025 | L | M | M | L | L | M | M |
| Alyabsi, 2022 | M | M | M | L | H | H | H |
| Dailey, 2013 | L | M | M | M | H | H | H |
| Konda, 2020 | H | M | M | M | M | M | H |
| Long, 2022 | L | M | M | M | L | L | M |
| Murphy, 2008 | H | L | M | M | H | H | H |
| Pinto, 2023 | M | ? | M | M | H | M | H |
| Rahman, 2024 | M | ? | M | M | M | H | H |
| Tang, 2023 | L | L | M | M | M | H | H |
| **Any/major complications** | | | | | | | |
| de Vries, 2020 | M | M | M | L | H | M | H |
| Roth, 2025 | M | M | M | M | H | M | H |
| Stenberg, 2019 | L | M | M | L | H | L | H |
| Tang, 2023 | L | L | M | M | M | H | H |

L, low; M, moderate; H, high; ?, no information

Traffic-light display of the six QUIPS domains plus the result-level overall judgment for every synthesized result in the five manuscript families (46 results from 39 unique studies; the broad survival family shown includes the 9 results of the primary postoperative all-cause survival analysis). A study contributing a result to more than one family is shown under each family; within a family, results are ordered alphabetically by first author. Columns: study participation; study attrition; prognostic factor (exposure) measurement; outcome measurement; study confounding; statistical analysis and reporting; result-level overall judgment. L, low; M, moderate; H, high; ?, no information; cell color duplicates the letter (green, low; yellow, moderate; red, high; gray, no information). The grid is split into two columns for page fit. Judgments are reproduced verbatim from the public frozen release; included studies without an analyzed estimate are not applicable. QUIPS, Quality In Prognosis Studies tool.

### eTable 9. Result-level risk of bias (QUIPS) summary

| Family | Results | High | Moderate | Low | High risk-of-bias share of random-effects weight |
|---|---|---|---|---|---|
| Postoperative survival (strict primary node) | 9 | 7 | 2 | 0 | 77.72% |
| Broad all-origin survival (sensitivity node) | 16 | 12 | 4 | 0 | 70.31% |
| Early postoperative mortality | 8 | 5 | 3 | 0 | 51.93% |
| Non-home discharge | 9 | 5 | 4 | 0 | 50.79% |
| Any or major complications | 4 | 4 | 0 | 0 | 100.0% |
| Healthcare-burden dispersion model (k=19; not an effect claim) | 19 | 12 | 7 | 0 | 56.33% |
| Unplanned readmission across all eligible windows | 9 | 7 | 2 | 0 | 63.16% |
| Prolonged length of stay (subfamily; not reported as an effect claim) | 4 | 4 | 0 | 0 | 100.0% |
| Resource use (subfamily; not reported as an effect claim) | 3 | 2 | 1 | 0 | 85.3% |

Result-level overall QUIPS judgments and the random-effects weight carried by high-risk results. The strict primary node is fitted as its own model; its weights are determined by the deposited standard errors and its between-unit variance and reproduce the high-risk share used in Table 2. The registry subfamily identical to non-home discharge is not repeated; length-of-stay and resource-use subfamilies were fitted only as dispersion diagnostics of the healthcare-burden family and are not reported as findings.

### eTable 10. Formal prognostic certainty (GRADE, Claim B) by family and domain

| Family | Risk of bias | Inconsistency | Indirectness | Imprecision | Publication bias | Certainty |
|---|---|---|---|---|---|---|
| Postoperative survival (strict primary node) | serious (-1) | serious (-1) | not serious (0) | serious (-1) | not assessed (0) | Very low |
| Broad all-origin survival (sensitivity node) | serious (-1) | serious (-1) | not serious (0) | serious (-1) | not serious (0) | Very low |
| Early postoperative mortality | serious (-1) | serious (-1) | not serious (0) | not serious (0) | not assessed (0) | Low |
| Non-home discharge | serious (-1) | serious (-1) | not serious (0) | not serious (0) | not assessed (0) | Low |
| Any or major complications | very serious (-2) | not serious (0) | not serious (0) | serious (-1) | not assessed (0) | Very low |
| Healthcare-burden dispersion model (k=19; not an effect claim) | serious (-1) | serious (-1) | serious (-1) | not serious (0) | not serious (0) | Not a single effect claim |
| Unplanned readmission across all eligible windows | serious (-1) | not serious (0) | not serious (0) | serious (-1) | not assessed (0) | Low |
| Prolonged length of stay (subfamily; not reported as an effect claim) | very serious (-2) | serious (-1) | not serious (0) | serious (-1) | not assessed (0) | Very low |
| Index length of stay (k=1; not fitted) | Not assigned | Not assigned | Not assigned | Not assigned | Not assigned | Not assigned |
| Resource use (subfamily; not reported as an effect claim) | very serious (-2) | serious (-1) | not serious (0) | serious (-1) | not assessed (0) | Very low |

Prognostic-factor GRADE starting at high, without automatic downgrading for observational design; domain judgments and totals are reproduced from the frozen certainty registry. Indirectness was rated not serious wherever exposure and outcome matched the registered definitions; the concern that discharge destination depends partly on the resources defining the exposure is stated in the Results as an interpretive caution, not as a downgrade. Claim F (that changing social connection would change the outcome) is not directly evaluated for any family. Grades apply to pooled effect claims; evidence-map completeness is descriptive and not graded.

### eTable 11. Small-study-effect diagnostics

| Analysis | k | Egger P value |
|---|---|---|
| Broad all-origin survival (sensitivity node) | 16 | .14 |
| Healthcare-burden dispersion diagnostics (not an effect claim) | 19 | .005 |

Performed only where at least 10 analysis weight units were available; exploratory. Statistical asymmetry neither establishes nor excludes publication bias, and its absence does not exclude it. Diagnostics for superseded earlier-generation analyses are retained in the repository technical appendix, not here.

### eFigure 4. Contour-enhanced funnel plots (k ≥ 10 families)

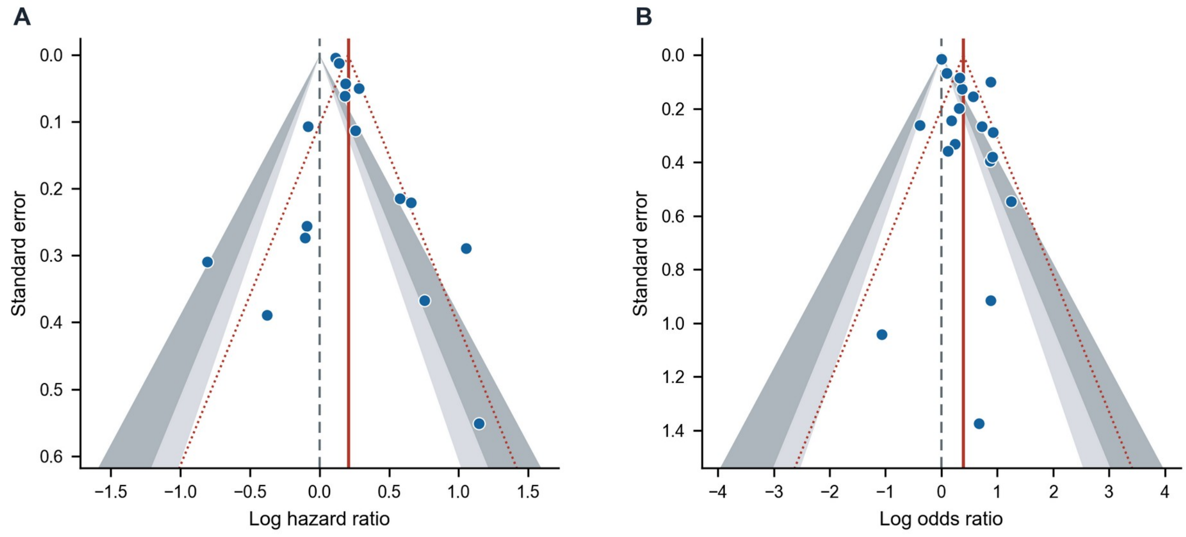


A, Broad all-origin survival (sensitivity node; k = 16; Egger *P* = .14). B, Pooled healthcare-burden dispersion model (k = 19; Egger *P* = .005; not presented as a single effect claim). Points are the locked per-unit log effects and standard errors; solid red line, locked REML pooled estimate; dotted red lines, pseudo-95% funnel bounds; dashed gray vertical line, the null (log ratio 0). Shading marks contour-enhanced significance bands relative to the null: light gray, .05 < *P* < .10; dark gray, .01 < *P* < .05; unshaded, *P* > .10 (center) and *P* < .01 (outer). Principal analyses (k = 4-9) are not funnel-assessed per the frozen statistical analysis plan; asymmetry neither establishes nor excludes publication bias. k, number of independent analysis weight units; REML, restricted maximum likelihood.

### eTable 12. Failure-to-rescue exposure-outcome cross-gap

Two independent search strategies (a full-text scan of the retrieved corpus and a structured scan of the estimate and study ledgers) identified 8 reports that genuinely analyzed or reported failure-to-rescue outcomes; 0 estimated the association with preoperative social connection. Failure-to-rescue outcomes were present in the surgical literature, but no eligible study estimated their association with preoperative social connection: an exposure-outcome cross gap, not an absent outcome.

### eTable 13. Clinical and review-process limitations

| Limitation | Consequence | Handling in this review |
|---|---|---|
| All contributing evidence is observational; residual confounding by socioeconomic position, functional status and comorbidity is likely | Average associations may be overestimated | Result-level QUIPS confounding judgments; GRADE downgrading |
| Marital status dominates exposure ascertainment | Coarse proxy for postoperative support resources | Construct axis kept unpooled; construct mix reported per family |
| High-risk-of-bias results carry 50-100% of random-effects weight in every principal analysis | Certainty capped at low/very low | Weight shares reported; formal GRADE |
| Registry and nested cohorts share patients | Study counts overstate independent evidence; totals would double-count | Dependence clusters; k = analysis weight units; no aggregate participant total reported |
| Readmission windows differ across studies; no exactly defined window shared by >1 independent unit | Strict-window question unanswerable | Reported as evidence gap, not a null result |
| Publication-bias testing possible only for the two largest analyses, exploratory only | Reporting bias cannot be excluded | SAP k>=10 rule applied uniformly |
| Human confirmation verified source fidelity of analyzed records; it was not blinded independent dual review | Reviewer agreement does not evidence independent convergence | Process named and bounded explicitly in eMethods 4 |

### eAppendix 1. Data and code availability

A frozen deposit package contains the analysis registry, analysis membership tables, dependence-cluster registry, result-level risk-of-bias assessments, GRADE evidence profiles, sensitivity analyses (fixed-effect, leave-one-out, multiverse), figure source data, the human-confirmation summary, the frozen protocols and the deterministic analysis code (provenance identifier SR-49523c19b885c87a). The package is publicly deposited at github.com/yinchuan123/social-connection-surgery-meta (Zenodo DOI 10.5281/zenodo.22739144). No copyrighted article full texts are redistributed.

### eAppendix 2. Artificial intelligence: directive and prompt sequence

This appendix documents the instructions under which large-language-model agents operated in this review, their sequence, their dates and their revisions. eMethods 3 describes what the agents did; this appendix describes what they were told.

A. What an instruction consisted of. No agent decision was governed by free-form conversation. Every agent action was determined by three layers: (1) a hash-frozen specification document — the eligibility contract, the outcome and exposure vocabularies, the estimate-selection hierarchy, the dependence-clustering protocol, and the risk-of-bias and certainty protocols — frozen before the step it governs, except that screening began under the registered eligibility criteria, issued as a written instruction package, before the hash-locked specification (v1.0) was fixed on July 29, 2026; (2) a numbered written investigator directive, identified by a directive code (for example PI-877R-SCIENTIFIC-RELEASE-PATCH-01), which instantiated a specification, adjudicated an ambiguity in it, or authorized a build; and (3) a per-record payload, namely the bibliographic record or the full text of the report being assessed.

Layers 1 and 2 determine every reported decision and are reproduced or referenced below. Layer 3 is not reproduced: it consists of published articles under copyright, each identified in the reference list and, for every analyzed estimate, by a structured source locator giving the exact printed location of the extracted value. A reader with access to the same articles can reconstruct any instruction in full by combining layers 1 and 2 with the named source. An agent could not revise a frozen rule; outputs were revised only by issuing a further numbered directive.

Verbatim interaction logs are not deposited. Beyond the copyrighted full texts entered into them, they contain author contact and administrative material unrelated to the review. The numbered directives and prompt templates are retained in the project archive and are available from the corresponding authors. Their volume and per-model breakdown are reported in the repository technical appendix.

B. Frozen specifications in force.

| Specification | Governs | Location | SHA256 |
|---|---|---|---|
| Eligibility criteria specification (locked v1.0) | Screening and full-text eligibility | Public deposit, protocol directory | sha256:8a6d5185b70e6575 |
| Formal prognostic GRADE protocol (v1.1) | Certainty of evidence | Public deposit, root | sha256:847f9265fa5d1494 |
| Signed scientific release SR-49523c19b885c87a | Deterministic analysis code and authoritative registries | Public deposit, release manifest | sha256:fdc9b96600cc5cfc |

C. Sequence of phases, with dates. Each phase was closed before the next was opened; no phase reopened a frozen artifact of an earlier phase except through a numbered directive. Where a phase could not be dated to the day from a frozen artifact, the window shown is bounded by the first logged session and the signed release, and the bounding evidence is named.

| Phase | Dates | Standing instruction to the agent | Governing frozen specification | Human act that closed the phase |
|---|---|---|---|---|
| 1. Search and deduplication | 2026-07-16 to 2026-07-17 | Not an agent task. Database searches were executed and records exported; agents performed deduplication bookkeeping only. | Search strategies, reproduced verbatim per database (eMethods 1) | Last-search date fixed at 2026-07-17; no database searched thereafter (the search-date reconciliation record) |
| 2. Title and abstract screening | from 2026-07-18 | Judge each record against the eligibility contract and return a coded decision with the clause relied on. Two agents judged independently and blinded to each other; a third adjudicated every disagreement. | The registered eligibility criteria, issued to the screening agents as a written instruction package; the hash-locked eligibility specification (v1.0) was fixed on 2026-07-29, during the review (public deposit, protocol directory) | Investigator rulings on protocol ambiguities, issued as numbered directives |
| 3. Full-text assessment and retrieval | through 2026-08-05 | Assess each retrieved full text against the same contract; a separate verifier agent, holding the full text, re-judged a risk-weighted sample seeking | The eligibility criteria specification (locked v1.0, 2026-07-29; public deposit, protocol directory) | Full-text retrieval closure recorded 2026-08-05 (the frozen PRISMA count record) |

| | | | | |
|---|---|---|---|---|
| | | grounds to overturn the decision. | | |
| 4. Corpus closure (identity gap wave) | 2026-08-04 | Finalise the 28 already-retrieved records still carrying non-terminal states. No new search; within-flow reclassification only. | Corpus closure protocol; PRISMA counts unchanged | PI approval of the material operational clarification, 2026-08-04 |
| 5. Structured extraction | 2026-07-18 to 2026-08-31 (bounded by the first logged session and the signed release) | Extract only values printed in the source into the controlled vocabulary, attaching a verbatim source locator to every analyzed estimate. Never infer, never auto-fill; where a value is not printed, record NOT_REPORTED. | EVIDENCE_REPRESENTATION_CONTRACT_v2; ADJUSTMENT_TWO_AXIS_SPEC_v1 | Third-pass adjudication from source where the dual pass disagreed |
| 6. Dependence adjudication and clustering | 2026-07-18 to 2026-08-31 (bounded by the first logged session and the signed release) | Cluster cohort entities that share participants under the registry-provenance rules; at most one weight unit per cluster per analysis. | COHORT_OVERLAP_RESOLUTION_PROTOCOL_v1 | PI directives established two cluster edges the rules could not settle |
| 7. Estimate selection | 2026-07-18 to 2026-08-31 (bounded by the first logged session and the signed release) | Where several eligible estimates exist for one outcome, select one through the frozen source-based hierarchy. The hierarchy may never use effect direction, interval width or null-crossing; re-run every legal alternative as a multiverse. | ESTIMATE_SELECTION_PROTOCOL; PAPER1_ESTIMATE_SELECTION_PROTOCOL_v1 | PI adjudicated level semantics and ties |
| 8. Risk of bias and certainty | 2026-07-18 to 2026-08-31 (bounded by the first logged session and the signed release) | Judge QUIPS at the level of the result, not the study; derive result-level judgments from the study level by frozen rule; compute certainty from the frozen GRADE protocol rather than rating de novo. | The formal prognostic GRADE protocol, version 1.1 (public deposit) | PI adjudicated the domain criteria and froze the protocol before application |
| 9. Synthesis | 2026-07-18 to 2026-08-31 (bounded by the first logged session and the signed release) | Fit only what the frozen analysis plan permits; run leave-one-out and the selection multiverse for every principal analysis; never pool across effect-measure types. | Frozen statistical analysis plan (eMethods 2) | Independent clean-room statistical reproduction of all locked results |
| 10. Two-person source confirmation | 2026-08-31 | Agents generated source-grounded recommendations only; the confirmation decisions themselves were not delegated. | FINAL_HUMAN_CONFIRMATION_METHODS | All 39 principal-analysis source records confirmed against the printed sources by Chuan Yin and Zehao Jing, separately; signatures verified against the archived signed document |
| 11. Release authorisation | 2026-08-31 | Compile the release and run every gate; a build may not be promoted while any gate fails. | the release manifest | PI signature of scientific release SR-49523c19b885c87a |

| | | | | |
|---|---|---|---|---|
| 12. Analyzed-size sign-off | 2026-09-05 | Revisit sizes not established at extraction; one agent re-derived each, a second re-derived it independently; disagreements were escalated, never resolved by the agents. | Analyzed-size protocol (eMethods 5) | 18 sizes and 10 not-printed findings checked against the printed source and signed by 2 investigators (0 corrections); the 9 bounding sizes recovered in the second pass and the remaining 18 cohort totals were not human-checked |
| 13. Manuscript preparation | 2026-08-31 to 2026-09-09 | Draft and revise from the signed release only; zero recomputation; every reported number must resolve to a frozen ledger field. Reference metadata may not be generated by the model. | Signed release SR-49523c19b885c87a | Authors reviewed all generated content and accept responsibility for it |

D. Automation and human governance boundary, reproduced from the frozen governance table of the signed release with internal notation removed; no wording is otherwise changed.

| Stage | AutomatedComponent | HumanComponent | Control |
|---|---|---|---|
| Screening and eligibility | system | PI spot adjudication on escalation | eligibility contract frozen before screening |
| Extraction of printed values | system (dual pass) | third-pass adjudication from source | precedence contract; no auto-fill |
| Identity and dependence clustering | system | PI directives established two cluster edges | one weight per cluster per analysis |
| Estimate preselection | system (13-level hierarchy) | PI adjudicated level semantics and ties | a level may discriminate only when every candidate is judgeable |
| Statistical estimation | system (deterministic) | none | independent clean-room reproduction |
| Risk of bias and certainty | system (result level) | PI adjudicated domain criteria and froze the protocol | no formal GRADE below the family level |
| Abstention decisions | system | PI reviewed each abstention | thresholds prespecified; abstention leaves a visible row |
| Escalation of ambiguous decisions | system raises, never decides | PI decides | escalation card records both options and their measured consequences |
| Principal source confirmation | system recommends | two people confirm | recommendation-assisted, not blinded independent review |
| Release authorisation | system compiles and gates | PI signs | signature archived with SHA-256 |

E. Models and dates of use, computed at build time from the execution logs. Interaction volume is reported in the repository technical appendix.

| Model | First | Last |
|---|---|---|
| claude-fable-5 | 2026-07-18 | 2026-09-02 |
| claude-opus-4-8 | 2026-07-18 | 2026-08-12 |
| claude-sonnet-5 | 2026-07-21 | 2026-07-25 |
| claude-opus-5 | 2026-07-27 | 2026-09-09 |
| claude-fable-5-1 | 2026-09-02 | 2026-09-13 |

Overall period 2026-07-18 to 2026-09-13. The runtime build identifier of the agent was not retained and is disclosed as a reproducibility limitation (eMethods 3). Reproduction of every reported result depends on the frozen ledgers, source locators and deterministic code in the deposit, not on re-running the agents.

F. Revisions. Revision was a controlled act, not a conversational one. A frozen specification could be changed only by a numbered directive that recorded the change and its consequences; the resulting departures from the registered protocol are itemized, with dates, in eTable 1, and the analysis-level history in the protocol and amendment index. Where a specification changed after evidence had been screened under the previous rule, the affected records were re-judged rather than carried forward.

G. Tasks never delegated to an agent: reference generation and reference formatting (all reference metadata were retrieved from PubMed and Crossref records); authorship decisions; the two-person confirmation decisions themselves; and signature of the scientific release.

This supplementary material has been provided by the authors to give readers additional information about their work.
